# Multimodal axes reveal individualized amyloid-β, tau, and neurodegeneration coupling in aging and Alzheimer’s disease

**DOI:** 10.64898/2026.05.24.26353955

**Authors:** Konstantinos Poulakis, Konstantinos Ioannou, Gleb Bezgin, Konstantinos Chiotis, Yasser Iturria-Medina

**Author notes:** Corresponding author: Konstantinos Poulakis **Adress and email:** Konstantinos Poulakis, PhD | Department of Neurobiology, Care Sciences and Society | Karolinska Institute | Center for Alzheimer’s research | Division of Clinical Geriatrics| Neo, floor 7, Blickagången 16, 141 52 Huddinge, Sweden |. **Author Contributions:** Paste the author contributions here.

## Abstract

Can we decode Alzheimer’s disease (AD) heterogeneity into a few portable axes that capture how amyloid-β, tau and neurodegeneration (A-T-N) spatially co vary in vivo? To answer this question, we built a pipeline that harmonizes longitudinal amyloid-β/tau PET and T1 MRI (gray matter) from ADNI cohort (12,430 images) with mixed effects modeling and then derived stage specific multimodal axes (mVCs) using linked component analysis, with robustness tested in simulations and external validation in the OASIS cohort (4,958 images). We identified a small set of multimodal axes that (i) recapitulate early tau weighted variation in cognitively unimpaired (CU) individuals, AD like A -T-N coupling in cognitively impaired (CI) individuals and atypical CU and CI participants with posterior (precuneus/occipitoparietal) and fronto insular/frontal weighted patterns, (ii) map onto domain specific cognition, APOE e4, and blood/CSF biomarkers of neurodegeneration, neuroaxonal injury and astrocyte activation, (iii) predict clinical transitions, (iv) generalize in an independent cohort, and (v) demonstrate modelling robustness to missing data, high dimensionality, and cross-cohort variability, enabling direct application of the extracted axes to new datasets for biomarker discovery and stratification. Multimodal axes provide a portable, interpretable layer for quantifying amyloid-β-tau-neurodegeneration coupling at the individual level, complementing current biomarker-based staging frameworks based on A-T-N status and tau PET topography, and can be computed on new datasets to aid clinical assessment and trial enrichment.

**Significance Statement:** We developed and validated a multimodal statistical pipeline to identify individualized patterns of association among core Alzheimer’s disease biomarkers: amyloid-β deposition, tau accumulation, and neurodegeneration. Applied to longitudinal PET and MRI data, the approach revealed distinct, reproducible axes of biomarker coupling across cognitively unimpaired and impaired individuals, linked to cognitive performance and clinical progression. By providing subject -level scores that quantify how pathologies co-express across brain regions, this framework supports fine-grained biomarker discovery, improves interpretation of Alzheimer’s disease heterogeneity, and can be extended to high -dimensional multimodal datasets in future biomarker studies

## Introduction

Alzheimer’s disease rarely conforms to rigid discrete categories ^1^. In late life, amyloid-β, tau, and neurodegeneration frequently co-occur, progress at different rates, and exhibit overlapping yet partially independent brain topographies within the same individual. While subtype^23^ and event-based^4^ models have been invaluable for mapping dominant trajectories, they predominantly classify individuals into distinct progression groups. This categorization risks losing the continuous, structured overlap created by mixed pathologies^5^ and resilience mechanisms^6^, which ultimately shape clinical expression. Consequently, within contemporary biomarker-based frameworks, including A-T-N^7^ classification and tau PET-based staging by regional topography^8^, there remains a need for patient-level representations that quantify how biomarkers couple within individuals, rather than only whether each biomarker is abnormal. Such a representation would quantify whether amyloid-β, tau, and neurodegeneration are elevated in the same individuals and brain regions, and whether their spatial patterns vary together across people.

Although existing data-driven approaches have begun to address this heterogeneity, important gaps remain. Many models rely on a single modality ^2^, are limited to cross-sectional data^9^, or evaluate relationships between only two modalities at a time^10^. Crucially, few frameworks are designed to disentangle the variance shared across multiple modalities from the variance specific to each. As a result, they may underrepresent the graded, overlapping nature of A-T-N interactions across clinical stages. A multi-view strategy that integrates amyloid-β- and tau-PET with structural MRI, and explicitly separates joint, pairwise, and modality-specific contributions, is well positioned to characterize these interactions. In this context, we use “N” in A-T-(N) as a pragmatic, imaging-based proxy of neurodegeneration (gray-matter thickness/volume), acknowledging that neurodegeneration is broader than atrophy including hypometabolism and synaptic loss among others. Thus, a key unresolved challenge is how to derive patient-level, multimodal representations that preserve continuous overlaps among amyloid-β, tau, and neurodegeneration while remaining interpretable enough for biological and clinical use.

To address this gap, we developed a stage-tuned multimodal decomposition framework that represents heterogeneity as overlapping, continuous axes called multimodal variance components (mVCs), capturing how amyloid-β (A), tau (T), and brain atrophy (N) couple within individuals rather than assigning categorical labels. First, we modeled longitudinal amyloid-β-, tau-PET, and structural MRI trajectories using mixed-effects models to estimate subject-specific biomarker deviations at a common age reference, and we handled structured and random missing data. Then we used a structured multi-view decomposition model (SLIDE^11^) to derive continuous axes that separate joint, pairwise, and modality-specific components of A-T-(N) coupling. These anatomically interpretable axes come with individual scores, offering a compact language for “how biomarkers co-express” inside a person rather than assigning persons to a single category. Using large cognitively unimpaired (CU) and impaired (CI) cohorts, we then (i) mapped these axes to domain-specific longitudinal cognition, (ii) associated them to APOE e4 and blood/CSF markers of pathology and injury, (iii) related baseline axis expression to clinical transitions (CU→MCI/AD; MCI→AD) with cross-validated models, and (iv) tested portability by projecting to an independent cohort and fitting a de novo model.

Our ultimate aim is to complement contemporary biomarker-based frameworks, including A-T-N classification and tau PET topographic staging, with an interpretable layer that quantifies individualized amyloid-β-tau-neurodegeneration coupling and supports biomarker interpretation, risk stratification, and trial enrichment.

## Results

We first modeled longitudinal amyloid-β PET, tau PET, cortical thickness, and subcortical volume to estimate region of interest (ROI)-level subject-specific biomarker deviations at age 75 (chosen because it provided a clinically relevant anchor within the overlapping age range of the cohorts and diagnostic groups) and annual slopes for the ADNI (discovery) and OASIS (validation) cohorts. The population intercepts and slopes were retained to show clinical stage-specific biomarker patterns and anchor the mVC interpretation (Fig. S-1). The random intercepts underwent imputation-scaling and were analyzed with the structured multiview decomposition model for mVC estimation (see Online Methods-pipeline overview). Unless noted, Results report ROI-level subject specific intercepts from mixed-effects models; model details and diagnostics are in Online Methods.

### What baseline amyloid-β, tau, and atrophy patterns anchor the axes?

At baseline, CU participants showed relatively preserved medial temporal cortical thickness/volume with milder fronto-parietal/occipital thinning compared to CI (MCI/AD) that exhibited more pronounced temporal and lateral parietal atrophy, consistent with symptomatic AD (Fig. S-1). Age-related atrophy in CU was evident in frontotemporal, cingulate, and lateral occipital regions, while CI showed a typical AD-like atrophy pattern. In CU, amyloid-β was relatively higher at baseline in subcortical and parietal/cingulate ROIs, whereas tau concentrated in temporal/subcortical regions; longitudinally, tau increases were most apparent across temporoparietal and lateral frontal/occipital cortices. In CI, both amyloid-β and tau were diffusely elevated at baseline, with tau showing steeper increases than amyloid- β (vs CU). Clinical stage-specific cognitive profiles corresponding to these global A-T-(N) levels are summarized in Table S-1/-2.

### Do diagnosis-specific models recover stage-dependent variance?

We estimated mVCs separately in CU and CI clinical stages and in a diagnosis-agnostic model (discovery cohort). The combined model recovered fewer clinical stage-specific patterns: only 0% (CU) and 5% (CI) of its leading mVCs closely matched the diagnosis-specific solutions (correlated with r > 0.9), indicating that pooling diagnoses obscures clinical stage-dependent variance. We therefore focused on CU-specific and CI-specific models. After pruning correlated solutions, 11 minimally redundant mVCs remained for downstream analyses.

### The multivariate architecture of amyloid-β, tau, and atrophy covariation in healthy and pathological aging

Each mVC is an axis of between-person variation; maps display −2 vs +2 SD contrasts (Fig 1A, Fig. 2A ; higher scores = stronger expression of that axis’ amyloid-β/tau/atrophy topography). In ADNI, 11 mVCs summarized A-T-(N) covariation; five “**typical**” axes (>10% data variance) captured most structure (Fig. 1A).

**Figure 1.**
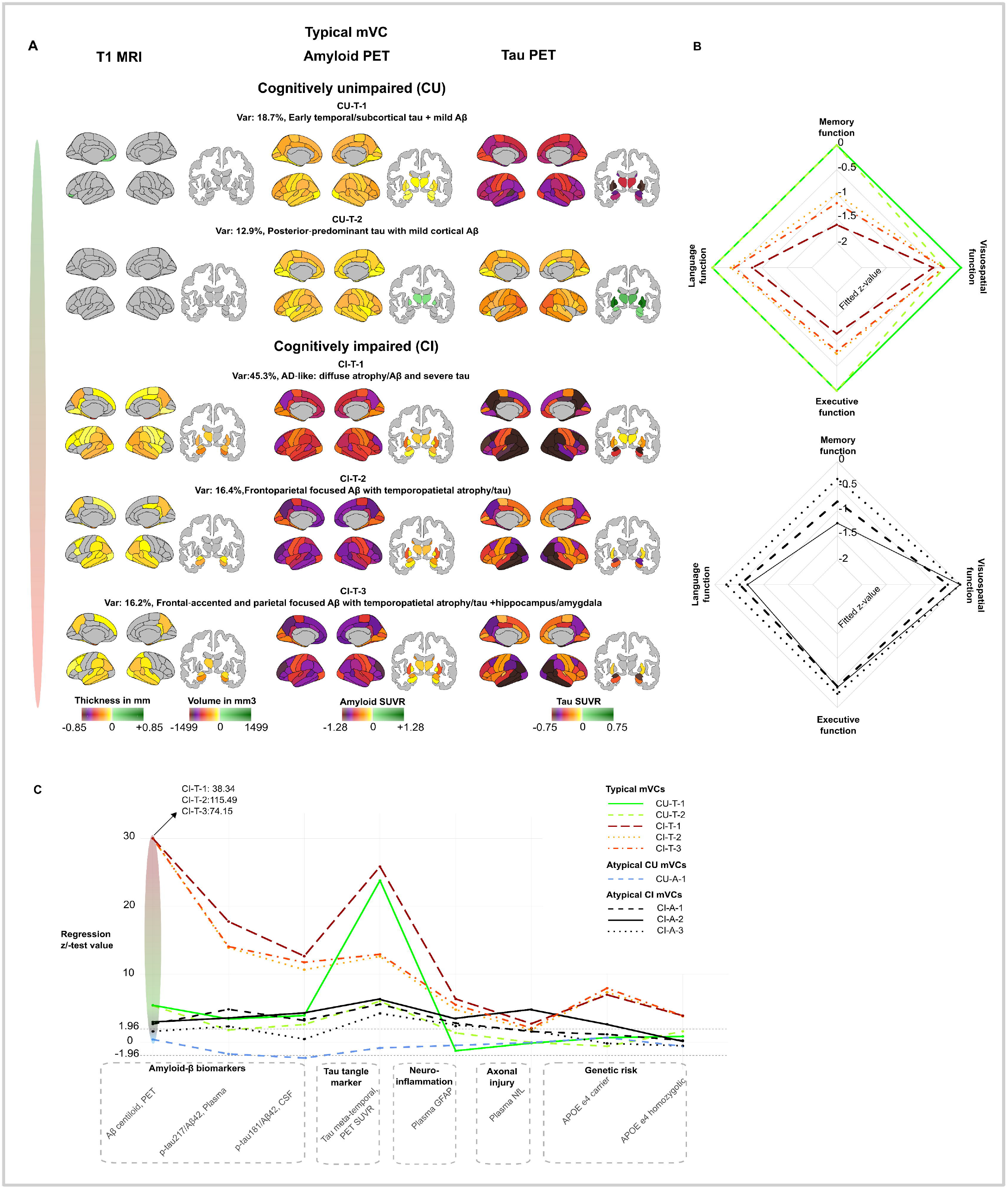
Typical CU/CI axes of biomarker variation, cognitive, and fluid/genetic associations. **A**. Shown are brain maps of multimodal variance components (mVCs) that capture the dominant axes of A-T-N(atrophy) coupling in cognitively unimpaired (CU; CU-T1, CU-T2) and cognitively impaired (CI; CI-T1, CI-T2, CI-T3) participants. For each axis, cortical thickness/volume (T1 MRI), amyloid-β-PET (SUVR), and tau PET (SUVR) are visualized as ROI-wise contrasts between individuals at −2 vs +2 SD on the mVC score (Methods, Eq. 8). non-significant ROIs are gray. Color bars reflect fixed truncation based on the CU random-intercept ranges to preserve contrast across mVCs (increasing pathology color: yellow, orange, red, purple, brown, black, maroon). Axes were estimated in stage-specific multiview models (separate CU and CI fits) following longitudinal mixed-effects harmonization (age anchored at 75 y; intercepts used for visualization) and SLIDE decomposition; the variance proxy noted per axis summarizes relative contribution to multimodal structure. Baseline CU and CI A-T-N(atrophy) profiles are shown elsewhere (Fig S-1). **B**. Radar plots summarize standardized baseline associations between each mVC and AD-relevant cognitive domains (memory, language, executive, visuospatial). Effects are z-scaled regression coefficients per +2 SD increase in mVC score from Gaussian mixed-effects models (age and education adjusted; Methods, Eq. 9). Lines denote point estimates; Typical CI axes (lowest fitted z-values in red/orange/yellow color) show the largest memory-led penalties, whereas CU-T-2 emphasizes visuospatial vulnerability. **C**. Lines show standardized effect sizes as z or t-values depending on regression model fitted corresponding to a shift from -2 to +2 SD in mVC score, estimated in age-adjusted models. Fluids include tau PET meta-temporal SUVR, amyloid- β centiloids, plasma p-tau217/Aβ42, CSF p-tau181/Aβ42, plasma NfL, and plasma GFAP (Gaussian link); APOE e4 carriership was modeled with logistic regression (logit link). Typical CI axes show the strongest alignment with ε4 enrichment, higher tau burden, and elevated NfL/GFAP, whereas CU-T axes exhibit early tau-weighted signatures with minimal atrophy. Brain maps, cognitive score and fluid genetic marker statistical comparisons were FDR-corrected (Benjamini–Hochberg, q<0.05). Aβ, amyloid-β; A- T-N(atrophy), amyloid-β-tau-neurodegeneration framework; CI, cognitively impaired (MCI/AD dementia); CU, cognitively unimpaired; FDR, false discovery rate; GFAP, glial fibrillary acidic protein; mVC, multimodal variance component; NfL, neurofilament light; ROI, region of interest; SD, standard deviation; SLIDE, structured latent factorization; SUVR, standardized uptake value ratio.

**Figure 2.**
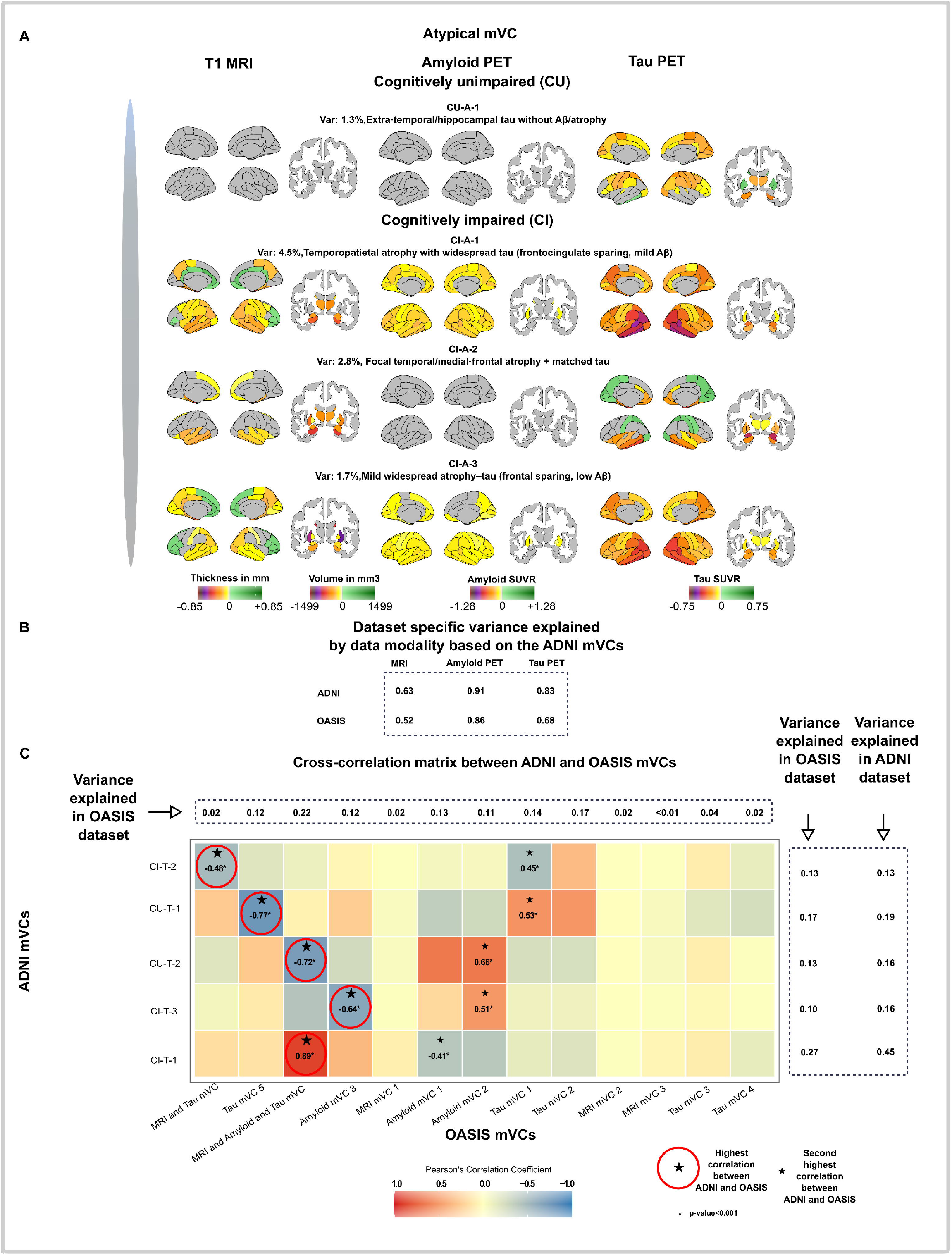
Atypical CU/CI axes of biomarker variation, cross-correlation between ADNI and OASIS biomarker patterns, and variance structure. **A**. Visualization matches Fig. 1A. **B**. Table entries report the median feature-level *R*^2^ within each modality (MRI, amyloid-β-PET, tau PET) computed from ADNI mVCs and applied to ADNI (in-sample) and OASIS (out-of-sample) data. For OASIS, subjects were projected onto ADNI mVCs using the learned mVC loadings. This quantifies how much of each modality’s signal structure is captured by ADNI axes in each cohort, demonstrating portability across datasets despite tracer/site differences harmonized in preprocessing. **C**. Heatmap shows Pearson’s correlation between the dominant ADNI axes (CU and CI models; rows) and OASIS axes (estimated de novo, columns). High correlations between selected ADNI-derived and de novo OASIS axes indicate cross-cohort concordance of dominant multimodal patterns. Right and top bars display the modality-aggregated variance proxy (median feature-level *R*^2^) for the corresponding axis in ADNI and OASIS, respectively, indicating that typical CI-like axes retain the highest variance contributions and cross-cohort concordance (CI-T-1 explains 45% variance in ADNI and 27% in OASIS), while low-variance axes show the expected attenuation. Aβ, amyloid-β; ADNI, Alzheimer’s Disease Neuroimaging Initiative; CI, cognitively impaired; CU, cognitively unimpaired; mVC, multimodal variance component; OASIS, Open Access Series of Imaging Studies; ROI, region of interest; SD, standard deviation; SLIDE, structured latent factorization; SUVR, standardized uptake value ratio.

Two CU axes were tau-weighted, with regional tau variation contributing more prominently than amyloid- β or atrophy variation, while amyloid-β variability remained present in regions relevant to early amyloid accumulation. **CU-T-1** showed a temporal-dominant emphasis, whereas **CU-T-2** showed a posterior cortical emphasis. Three CI axes (**CI-T-1/-2/-3**) were AD-like: higher scores combined temporoparietal/precuneus atrophy, advanced neocortical tau, and diffuse cortical amyloid-β (**CI-T-1** showed the greatest variance in pathology and subtle frontal/limbic shifts distinguished **CI-T-2/-3**). Axes quantify coupling strength between biomarkers along recognizable routes rather than discrete stages.

Lower-variance (“**atypical**”, 1–10% data variance) axes captured region-selective patterns (Fig. 2A). In CU, **CU-A-1** reflects isolated extra-temporal/hippocampal tau with comparatively weaker amyloid-β and atrophy contributions. In CI, **CI-A-1** represents severe temporoparietal atrophy with widespread tau, most prominent laterally in temporoparietal regions and detectable cortical amyloid-β variation; higher scores intensify both atrophy and tau. **CI-A-2** shows localized severe temporal and medial frontal atrophy accompanied by regionally matched tau; increasing scores accentuate these focal deficits without matching variance in amyloid-β. **CI-A-3** captures a milder widespread atrophy-tau pattern with relative frontal sparing, comparatively weaker amyloid-β contribution, and overall lower tau signal at low scores, strengthening with higher scores.

Very **low-variance** (<1%) axes mostly reflect additional atrophy contrasts. Given their small contribution, we analyze them briefly and refer to Fig. S-2. **CU-L-1** differentiates frontocingulate vs posterior/subcortical atrophy. Finally, **CU-L-2** contrasts mild subcortical/insular atrophy with cingulate/frontoparietal atrophy.. Across all components, the variance-aligned interpretation applies uniformly: low mVC scores denote weak expression of the underlying biomarker pattern, whereas high scores denote strong expression of the same pathology axis.

### How do axis scores map to cognition?

We next examined whether biomarker severity and topography (individual expression of each mVC axis) predicted baseline and longitudinal cognitive performance based on the ADSP memory/language/executive/visuospatial cognitive domains. Because mVC scores quantify the degree to which each subject expresses a given variance-derived biomarker pattern, higher scores indicate proportionally greater A-T-(N) involvement. Among typical CU components, higher **CU-T-2** scores (posterior-predominant cortical tau) were significantly associated with worse baseline visuospatial performance (z = -0.34; Fig. 1B and Table S-3). In contrast, higher scores on typical CI components (increasing A-T-(N) burden) showed broad cognitive associations: **CI-T-1/-2/-3** were each linked to worse baseline performance across multiple domains, with the largest effects in memory (z = -1.62/- 0.99/-1.18 respectively), followed by executive (z = -1.16/-0.75/-0.81 respectively), then language (z = - 0.8/-0.37/-0.44 respectively) and visuospatial (z = -0.56/-0.36/-0.35 respectively) function.

Atypical components demonstrated similar overall cognitive relationships, but with a distinct hierarchy of affected domains. Higher atypical CU and CI scores predicted the greatest impairment in memory, followed by language, then executive and visuospatial abilities, mirroring their A-T-(N) burden (Fig. 1B and Table S-3). Finally, low-variance components were associated with more selective non-memory deficits. In CU, lower **CU-L-2** scores, indicating greater insular-subcortical atrophy with posterior and cingulate tau were associated with worse baseline language function (z = +0.25). These mVCs were also associated with modest longitudinal decline, showing greatest effects in memory (CU-T-2/CI-T-1/ CI- A-2) and executive (CI-A-2) function (Table S-3). Further cognitive phenotyping of the mVC axes based on the MoCA (Table S-3) and Alzheimer’s Disease Assessment Scale (Table S-4) cognitive domains batteries were estimated and aligned with the results in the ADSP z-scores.

### Biological determinants revealed by amyloid-β, tau, and atrophy axes: genetic, CSF, and plasma correlates

To deeper understand the biological drivers of the mVC axes, we assessed their correlation with genetic, CSF and plasma markers of AD, neurodegeneration and inflammation (Fig. 1C, Table S-5/-6).

The apolipoprotein (APOE) genetic risk is associated largely with the typical CI components. The greatest association of heterozygote APOE e4 carriership was observed between the **CI-T-1** (odds of being APOE e4 positive compared to negative increase 50-fold by shifting from -2 to +2 SD in mVC score), **CI-T-3** mVCs (same as CI-T-1), and **CI-T-2** (odds increase 33-fold), followed by the low variance mVC **CU-L-2** (odd increase 5-fold) and finally the atypical CI component **CI-A-2** (odd increase ∼3-fold). The same order of relationship is maintained with APOE e4 homozygosity, where **CI-T-3**, is followed by **CI-T-1**, and finally **CI-T-2** indicating that typical patterns of AD pathophysiology are associated with the APOE e4 genotype. The estimated Tau PET meta-temporal SUVR and amyloid-β PET centiloid values’ association with mVCs follow closely the tau and amyloid-β burden with effects (Table S-5) proportional to PET binding intensity in the visualized mVCs (Fig. 2-A/-C).

Moreover, the novel blood-based AD biomarker p-tau217/Aβ42 correlate tightly with the typical CI mVCs, and to a lesser extent with the atypical CI mVCs and **CU-T-1** (Fig. 1C, Table S-5/-6). On the other hand, the CSF AD biomarker p-tau181/Aβ42 has an increasing correlation starting with negative values for the **CU-A-1**, followed by the Typical CU mVCs, then the Atypical CI mVCs, while highest associations are observed with typical CI mVCs showcasing a greater sensitivity to the neuroimaging biomarkers, at least for the tau pathology, compared with the plasma p-tau217/Aβ42. Plasma neurofilament light chain (NfL) correlated significantly with the typical **CI-T-1** and the atypical **CI-A-2** mVC axes, with the atypical mVC (CI-A-2) having 59% higher effect than the typical (CI-T-1) (Fig. 1C, Table S-5). Moreover, the **CI-A-2** axis showed one of the lowest amyloid-β centiloid burden increases by shifting from -2 to +2 SD in mVC scores (Table S-5/-6). Finally, plasma glial fibrillary acidic protein (GFAP) levels associated mostly with the typical and to a less extent with the atypical CI mVC scores. GFAP also correlated to a lesser extent with **CU-L-1** (progressive posterior and subcortical atrophy), one of the two CU low variance components.

### Do amyloid-β, tau, and atrophy axes scores predict clinical transitions?

Using a 5-fold cross-validated multinomial logistic model, we quantified how changes in mVC scores (−2 to +2 SD) relate to diagnostic transitions. Odds ratios (ORs) below 1 indicate that higher mVC scores are associated with a greater probability of progressing to a more impaired diagnostic category, whereas ORs above 1 indicate a tendency to remain in the same diagnostic stage rather than progress or regress. Among cognitively unimpaired (CU) individuals, the tau weighted **CU-T-2** component yielded very low ORs for CU→MCI (0.035) and CU→AD (0.001), indicating that higher **CU-T-2** scores markedly increase the likelihood of clinical worsening (Table S-7). **CI-T-1** showed a similar pattern (CU→MCI: 0.065; CU→AD: 0.001), reinforcing the role of temporal/tau signal in early decline. In contrast, **CU-L-2**, a low-variance MRI component, showed high ORs (CU→MCI: 25.119; CU→AD: 8.594), suggesting that increasing **CU-L-2** is linked to CU stability rather than progression. Notably, **CU-A-1** displayed an extremely high OR for CU→AD (∼3656), consistent with a pattern that is highly unlikely for AD-type progression, whereas **CI-A-2** showed a near-zero OR (∼0.00) for CU→AD, highlighting a predominantly temporal atrophy-tau signature closely tied to direct CU→AD transition. Amyloid-β-weighted components (**CI-T-2, CI-T-3**) also trended toward lower ORs for CU→MCI/AD, albeit less strongly than tau-dominated patterns.

In MCI-anchored contrasts, most components exhibited ORs >1 for MCI→CU, indicating that increasing mVC scores generally align with stability at MCI rather than clinical regression to CU. This was most evident for **CU-T-2** (80.53) and **CI-T-1** (44.48); **CU-T-1** was also high (17.308). For MCI→AD, nearly all available components showed ORs <1 (e.g., **CU-T-1**: 0.044; **CU-T-2**: 0.062; **CI-T-1**: 0.029; **CI-T-2**: 0.078; **CI-T-3**: 0.073; **CI-A-1**: 0.096; **CI-A-2**: 0.161), indicating that higher component expression, often reflecting tau, amyloid-β, and/or structural atrophy, corresponds to a greater probability of progression to AD compared with MCI stability. Together, these cross-validated ORs (adjusted for false discovery rate) highlight tau and temporal atrophy-related mVCs as robust markers of clinical decline, while certain atypical or low-variance patterns align more with diagnostic stability.

### Amyloid-β, tau, and atrophy covariance evaluated in the OASIS dataset

To test generalizability, we first projected OASIS participants onto the ADNI-derived mVCs and, in parallel, fit the multiview pipeline de novo in OASIS. Both approaches converged on AD-like axes as the dominant correlates of cognitive decline. In OASIS, projecting individuals onto ADNI-derived axes explained substantial out-of-sample variance (MRI ∼52%, amyloid-β∼86%, tau ∼68%; Fig. 2B) and preserved the AD-like rank order (**CU-T-1/-2**; **CI-T-1/-2/-3**; Fig. 2C). Differences in explained variance across modalities should be interpreted as properties of the learned representation and measurement structure, rather than direct evidence that one pathology is biologically more important than another. The typical CI axes again showed the largest memory penalties (±2 SD contrast), with CU-typical tau axes relating to memory mainly within CI, indicating that higher joint amyloid-β-tau-atrophy burden generalizes to worse memory in both impaired and unimpaired individuals (Table S-8). Moreover, **CU-T-1/-2** were associated with memory decline (MoCA Memory), but only within the CI stratum (−0.54), consistent with early tau vulnerability that becomes clinically evident after impairment. Selected atypical axes were related to domain-specific decline, memory for **CI-A-2** (−0.31, CU stratum). A de novo OASIS multiview fit recovered convergent AD-like axes, several correlating highly with ADNI CU/CI axes (Fig. 2C), with cognitive effects strongest for CI (Fig. S-9).

### Simulation-defined operating regime of the multimodal pipeline

Simulations designed to mirror our PET/MRI setting and evaluate more complex multimodal settings, showed that the SLIDE multiview decomposition, when integrated to our pipeline, recovered joint, pairwise, and modality-specific components across 50-5,000 features and 2-7 modalities, and remained accurate under random (≤30%) and structured missingness (Table S-10/-11); performance degraded only with very low per-modality dimensionality or when the number of modalities exceeded eight datasets (Fig. S-3). These results justify applying the framework to the three-modality ADNI/OASIS analyses and projecting axes across cohorts, while bounding interpretation to this validated operating regime; it also justifies the addition of more modalities in future studies. Full design and metrics (including cluster recovery r > 0.94–0.97) are in Methods and Fig. S-3 to S-6, Tables S-10/-11.

## Discussion

Alzheimer’s disease rarely follows a single canonical sequence, and categorical subtypes can miss the structured co-occurrence of pathology within individuals. By representing heterogeneity as continuous axes of A-T-(N) coupling strength, our multiview framework complements A-T-N and tau-PET topographic staging by providing interpretable, progression-sensitive measures rather than rigid discrete categories. This viewpoint respects the fact that amyloid-β, tau, and atrophy frequently co-occur and progress at partially independent rates, and it builds on, but does not replace subtype and event -based modeling by making the overlap itself measurable. Conceptually, the axes offer a compact language for “how biomarkers travel together” within a person, which can be used alongside clinical staging and screening tools to guide interpretation and decision-making.

The clinical-stage biomarker maps and the multivariate architecture of amyloid-β, tau, and atrophy covariation together suggest a useful distinction between population-level predictability and individual - level divergence. At the population level, amyloid-β, tau, and atrophy exhibit structured dependencies, such that one biomarker can partially inform expectations about the others. However, the mVCs show that individuals can depart substantially from these average relationships: heterogeneity is expressed not only in overall biomarker burden, but also in where pathology concentrates and how strongly modalities co-express regionally. As a result, a single biomarker is rarely sufficient to infer an individual’s full amyloid-β-tau-neurodegeneration profile, either in severity or spatial distribution. These person - specific departures from population-average coupling may reflect selective vulnerability or resilience to pathology. In some individuals, axis-specific deviations may correspond to distinct clinical variants, whereas in others they may capture the co-expression of AD pathology with additional age-related co-pathologies. Although this interpretation remains hypothesis-generating, it provides a biologically grounded rationale for using multimodal axes as individualized readouts of disease organization, capturing heterogeneity in both severity and spatial topography that cannot be inferred from a single biomarker alone.

In CU individuals, the mVC framework appeared more sensitive to regional tau-topographic variation than to global amyloid-β burden when relating biomarker patterns to subtle cognitive differences, consistent with evidence linking tau PET topography and amyloid-β-tau interactions to clinical and cognitive variability in early Alzheimer’s disease^12,13^. This does not imply that amyloid-β is unimportant; rather, once amyloid-β-related processes are present, regional tau variation may provide a more proximal readout of domain-specific vulnerability and cognitive expression^12,13^. These early tau-topographic signals may support closer monitoring and enrichment strategies for secondary -prevention trials^14^. Because the CU axes generalized to an independent cohort and preserved their relative rank in explained variance, they likely represent reproducible early signals rather than cohort -specific artifacts, in line with prior work identifying reproducible tau trajectories ^15^.

In symptomatic disease (CI), three AD-like axes (CI-T-1/2/3) capture overlapping routes of the A→T→N cascade, with broad cognitive penalties led by memory, followed by executive, language, and visuospatial domains, consistent with evidence that neocortical tau and its spread track cognition and cortical thinning more closely than amyloid-β^12^. Longitudinal findings further indicate that amyloid-β positivity is permissive for neocortical tau spread and that tau accumulation rate tracks cognitive decline^16,17^. Although AD-like, these axes are not redundant: subtle limbic–frontal tilts and frontal/anterior shifts separate them, mirroring clinically meaningful spectrums inside “typical AD”. This approach preserves overlap that fixed subtype assignments can obscure while still enabling stratification by dominant vulnerability, an advantage when aligning patients to therapeutic mechanisms and domain-matched endpoints.

Lower-variance atypical axes aligned with network-selective phenotypes, with posterior-weighted patterns reminiscent of posterior cortical atrophy-like visuospatial vulnerability and frontal/insular patterns aligned with executive-language deficits^18^. Notably, we identify an axis where medial temporal predominant tau, atrophy, couples primarily with memory impairment, and the most elevated NfL among mVCs. Given that NfL is a non-specific marker of neuroaxonal injury, this profile may reflect mixed or limbic-predominant neurodegenerative processes co-occurring with AD pathology, rather than a purely canonical AD pathway^19^. For clinicians, these axes provide an interpretable framework for understanding amnestic versus non-amnestic risk profiles that standard AD-centric schemes may underemphasize, while also helping to identify individuals in whom mixed-pathology profiles may inform stratification for targeted pharmacological approaches.

Axis expression was prognostic. Tau-laden and temporal/posterior signatures tracked a higher likelihood of CU→MCI/AD and MCI→AD, whereas certain low-variance MRI axes aligned with relative stability, reinforcing the notion that amyloid-β is necessary but not sufficient, while tau distribution and burden are proxies to near-term decline^17^. This suggests immediate uses in both practice and for trial enrichment: (i) in CU, heightened posterior/temporal tau expression argues for closer follow-up and targeted secondary prevention; (ii) in CI, higher AD-like axis scores favor T+/T+N-targeting strategies, with endpoints prioritized to memory and then executive cognitive domains, where effects are largest. Utilizing the CI-A-2 pattern as a marker of progression risk in CU individuals demonstrates the ability of the multiview framework to uncover meaningful risk signatures based on multimodal biomarkers.

The axes are biologically anchored. APOE e4 enrichment, especially homozygosity in the typical CI axes mirrors evidence that ε4 potentiates Aβ-tau coupling, and higher neocortical amyloid-β with temporoparietal tau, even partly independent of amyloid-β in medial temporal regions^20,21^. Fluid biomarkers separated in expected ways: plasma/CSF p-tau (181/217 tracked permissive amyloid-β and early pathophysiology better than tau-PET topography^16,22^. GFAP associated most with amyloid-β-rich CI, consistent with astrocytic activation that precedes or accompanies tau spread^23,24^. In contrast, NfL was most informative for the atypical temporal tau-atrophy profile, where strong neurodegenerative involvement occurred without a commensurate amyloid-β increase; this pattern is consistent with NfL as a non-specific marker of neuroaxonal injury and may indicate mixed or age-related co-pathology rather than AD-specific progression alone^25,26^. Together with the tau-PET topographies and cognitive associations, these biological triangulations support the multimodal axes as meaningful indicators of disease activity rather than statistical conveniences.

Projecting OASIS individuals onto ADNI-derived axes reproduced the rank order of explained variance and the memory penalties for CI and several CU axes; a de novo OASIS fit recovered convergent AD-like axes with the strongest cognitive effects in CI. Across modalities, explained variance ranked amyloid-β PET > tau PET > MRI thickness/volume, consistent with prior work showing most heterogeneity in atrophy subtypes^2,27^, amyloid-β plateauing in CI but remaining heterogeneous in CU/MCI ^9,10^, and tau subtypes partially mirroring atrophy^15,28^. This portability argues against cohort-specific overfitting; as expected, lower-variance axes showed weaker cross-cohort correlations. Because OASIS is CU-rich (86% vs 58% in ADNI) and used a combined CU/CI model, axes were more CU-weighted, likely attenuating cognitive associations^13^.

Subtyping and event-based models have clarified AD heterogeneity, but often force a single subject clustering or order^2^. An axis-based representation respects the structured overlap seen within individuals (including mixed pathologies and resilience) and quantifies A-T-(N) coupling at the patient level, a feature recognized in neuropathological work on A-T-N heterogeneity^5^. Clinically, a stage-tuned readout is actionable: (i) CU: high posterior/temporal T-dominant expression supports visuospatial monitoring and eligibility for Aβ/early anti-tau prevention; CU individuals with a CI-A-2–like profile (early increased temporal tau + atrophy) may warrant closer follow-up and tailored monitoring frequency. (ii) CI: CI-T scores (A→T→N) help stratify for amyloid-β or T+N trials and may guide cognitive domain-matched outcomes (memory first, then executive/language). Because mVC scores track stage-relevant coupling and increase with clinical worsening, they offer progression-sensitive, biologically interpretable endpoints aligned with AT(N)/tau PET-based topographical staging^7,13^ and a continuous scale that aids powering, interim monitoring, and cross-arm comparisons in trials.

We tailor multimodal fusion to the realities of longitudinal PET/MRI by harmonizing data with mixed-effects models (accommodating irregular sampling and enabling intercept/slope estimates), handling both structured and unstructured missingness, and decomposing joint, pairwise, and modality-specific variance into few anatomically interpretable axes that yield individual-level scores directly mappable to ROIs and usable in risk models and potentially clinical trials. The approach builds on established multimodal fusion frameworks (Linked ICA/FLICA, JIVE), but delivers a clinically focused, longitudinal implementation suited to consortia-scale neuroimaging where incomplete, uneven acquisition is common^29–31^. Using the SLIDE implementation^32^ of structured multiview decomposition, simulations defined the operating regime, high dimensionality with moderate missingness in which component recovery is reliable, supporting confidence that the reported axes and cross -cohort projections reflect signal rather than artifacts. Boundary conditions clarify where interpretation should be cautious (see Online Methods). These results justify application in PET/MRI and support prospective, simulation-based power planning (e.g., ≥80% recovery) to determine minimal sample size and identifiable rank under planned modalities, missingness, and effect sizes. Thus, the method fits cross-sectional/longitudinal PET/MRI with heterogeneous coverage, producing fit -for-purpose axis scores for risk stratification and trials within the validated operating regime.

A subset of mVCs were near-collinear in loadings but not in participant-level scores, consistent with rotational freedom and sampling variability; several CU/CI components also showed moderate cross-stage correlations without equivalence, indicating partial reuse across clinical stages. To limit redundancy, we retained axes by objective variance and pruned correlated solutions using a prespecified post-hoc algorithm (see Online Methods); alternative de-redundancy strategies (e.g., variance-inflation-factoring, correlation-based screening, orthogonalization/Procrustes) could yield slightly different, yet similar mVC sets. Despite cross-cohort validation and mixed-effects harmonization, residual heterogeneity (e.g., tracer differences, tau-PET topographic staging ROI definitions) can affect spatial sensitivity. Co-pathologies (e.g., Lewy body disease, TDP-43) were not modeled, so some atypical axes may reflect network-selective co-pathology. Axes are associational, not causal; interventional studies should test whether lowering axis expression yields clinical benefit (e.g., tau-lowering, astrocyte-modulating strategies). For prognosis, we modeled CU→stable/MCI/AD and MCI→stable/CU/AD using stage-specific CU/CI models with cross-validation, and also a diagnosis-agnostic multiview decomposition model that produced similar but fewer stage-specific mVCs; given low agreement and the ability of stage-specific models to recover stage-proximal patterns, we prioritized the latter. Prognosis could not be validated in OASIS (low CI%), so we provide subject -level mVC scores and loadings for projection (Supplementary data: Subject-specific mVC scores & mVC loadings per dataset). Most heterogeneity reflects biomarker severity and stage-specific models tend to recover stage-closer patterns (CU, MCI, AD)^3,33–36^; model choice should reflect study goals, disease, and data. Neuropathologic confirmation remains outstanding. Although developed in AD, the pipeline is disease-agnostic and portable to other multiview neurodegenerative cohorts.

## Conclusion

In summary, axes, not discrete categories, explain clinical heterogeneity in AD. By complementing A-T-N and tau-PET topographic staging with a measure of coupling A-T-(N) strength, our pipeline yields stage-tuned axes of overlap among amyloid-β, tau, and neurodegeneration that can be computed on new multimodal datasets. These axes provide a simple, interpretable layer that researchers and clinicians can use to enrich, stratify, and interpret their own data, making our approach well-suited for future analyses and trials as a fit-for-purpose component alongside existing methods.

## Materials and Methods

To comprehensively characterize disease-related variance across molecular and imaging biomarkers, we developed an unsupervised multi-view, multi-modal latent modeling pipeline for Alzheimer’s disease (AD) pattern discovery. The framework integrates complementary biomarker modalities, amyloid-β-PET, tau PET, and structural MRI within a unified decomposition model that jointly captures shared (joint), pairwise and modality-specific sources of variance. Designed for high-dimensional neuroimaging data, it balances analytical complexity with interpretability, ensuring that latent components remain directly traceable to their biological correlates. The approach enables robust estimation under incomplete data conditions, as validated on simulated datasets comprising up to seven modalities with 5,000 features each and up to 30% missingness. Applied to the ADNI and OASIS cohorts, the model delineated a dominant Alzheimer’s-related latent factor explaining the majority of multimodal variance, along with secondary modality-specific components reflecting distinct amyloid-β, tau, and grey matter anatomical trajectories in cognitively impaired (CI) and cognitively healthy (CU) aging. These latent signatures revealed convergent and divergent biomarker-cognitive relationships, identified clinically meaningful participant clusters, and mapped stage-specific neurobiological patterns across the AD continuum.

### Data

#### Cohorts and participants

The Alzheimer’s Disease Neuroimaging Initiative (ADNI) is a large, longitudinal multi-center study launched in 2004 to identify and validate biomarkers for early detection and disease tracking in Alzheimer’s disease (AD). It integrates multi-modal imaging (structural MRI, FDG-, amyloid-β-and tau-PET), genetics, cerebrospinal fluid biomarkers, and neuropsychological assessments. Participants include CU individuals, those with mild cognitive impairment (MCI), and patients with AD type dementia, diagnosed using standardized clinical and neuropsychological criteria such as the NINCDS-ADRDA guidelines and Clinical Dementia Rating (CDR) scores (https://adni.loni.usc.edu/). The Open Access Series of Imaging Studies (OASIS) provides publicly available cross-sectional and longitudinal brain imaging datasets to promote open neuroimaging research on aging and dementia (https://sites.wustl.edu/oasisbrains/). OASIS cohorts include cognitively normal older adults and individuals with very mild to moderate dementia due to AD, diagnosed using the CDR scale and clinical consensus procedures. Imaging data are standardized for structural analysis, enabling reproducible investigations of age-related and disease related brain changes.

#### Sample

We analyzed longitudinal data spanning healthy aging and the AD dementia spectrum, including cognitively unimpaired (CU) and impaired (CI) individuals. The discovery dataset included ADNI participants with CU, MCI, and AD dementia diagnoses. Replication was performed in the OASIS dataset with the same diagnostic categories. All participants had amyloid-β-PET and T1 MRI; longitudinal tau PET was required in ADNI and available cross-sectionally in OASIS. The final sample comprised **1**,**250 individuals and 8**,**850 images** (tau PET, amyloid-β-PET, and T1 MRI) across both cohorts. The study flowchart details the selection process (Figure S-7). For analysis, MCI and AD dementia were merged into a single CI group. Diagnosis was anchored to the closest clinical assessment at age 75. In total, 829 participants (477 CU, 352 CI) from ADNI and 421 participants (363 CU, 58 CI) from OASIS were included for model training and validation (Figure S-7, Supplementary data: Race distribution).

#### T1 MRI sample

MRI data from ADNI (ADNI-1, ADNI-GO/2, ADNI-3) and OASIS included 3T MP-RAGE scans with isotropic voxel sizes of 1.0–1.2 mm were included (Figure S-7). ADNI images passed visual QC (motion, coverage) and were processed with the cross-sectional FreeSurfer pipeline (versions 5.1/3 at UCSF/SF VA). OASIS images were processed with FreeSurfer 5.3 (2012, human connectome patch). We extracted mean thickness and volume for 68 cortical regions (Desikan–Killiany atlas) and 14 subcortical regions (hippocampus, amygdala, nucleus accumbens, thalamus, caudate, putamen, pallidum). Outputs were QC’d for segmentation errors; manual edits were performed in TkMedit when needed (http://freesurfer.net/fswiki/TkMedit). Cortical gray matter and intracranial volumes were also extracted. Full acquisition and processing details are available on ADNI (https://adni.loni.usc.edu/wp-content/themes/freshnews-dev-v2/documents/mri/Acquisition_Table.png ; https://adni.loni.usc.edu/wp-content/themes/freshnews-dev-v2/documents/mri/ADNI_MRI_overview_2.6.18.pdf) and the OASIS^37^ websites (https://sites.wustl.edu/oasisbrains/home/oasis-3/). Effects of FreeSurfer version and ADNI sub-study were statistically adjusted (see Analysis flowchart).

#### Amyloid-β PET sample

Amyloid-β PET assessed *Β* amyloid (A*Β*) deposits using florbetapir ([18F], FBP), florbetaben ([18F], FBB), and Pittsburgh Compound B ([11C], PiB). ADNI PET frames (FBP, FBB) were co-registered to the nearest T1 MRI and normalized to whole cerebellum uptake (UCSF/SF VA Medical Center, Amyloid-β-PET Processing Methods in https://adni.loni.usc.edu/). SUVRs were computed using the FreeSurfer Desikan–Killiany atlas. OASIS PET frames (PiB, FBB) were processed with the PET Unified Pipeline (PUP, https://github.com/ysu001/PUP), following similar steps: co-registration, intensity normalization, and atlas-based quantification. Centiloid (CL) values provided tracer-invariant amyloid-β burden estimates and defined positivity across cohorts. All segmentations were QC’d; failed outputs were excluded (Figure S-7). SUVRs from different tracers were harmonized to FBP-equivalent values (see Analysis flowchart).

#### Tau PET sample

Tau PET used flortaucipir ([18F] AV-1451) in both cohorts. Frames were co-registered to T1 MRI and smoothed to uniform resolution using the ADNI PET Core or OASIS PUP pipelines. SUVRs were computed using FreeSurfer segmentation and normalized to inferior cerebellar gray matter. ADNI defined a temporal meta-ROI from five bilateral regions (entorhinal, amygdala, fusiform, inferior temporal, middle temporal). OASIS used a similar composite (amygdala, entorhinal, inferior temporal, lateral occipital) with positivity threshold >1.22^38^ (see OASIS-3: imaging methods and data dictionary, version 2, July 2022). All outputs underwent manual QC; failed cases were excluded (Figure S-7).

#### Genetic, environmental, fluid (CSF and plasma) biomarkers, and cognitive data

We characterized cognitive function in ADNI and OASIS utilizing the Montreal Cognitive Assessment (MoCA) subtests and the Mini-Mental State Examination (MMSE) total score. For deeper phenotyping in ADNI, we leveraged 13 subtests from the Alzheimer’s Disease Assessment Scale (ADAS), along with composite scores capturing memory (ADNI-Mem), language (ADNI-Lan), executive (ADNI-Exe), and visuospatial (ADNI - Vsp) function^39^ (ADSP). In OASIS, we extended the cognitive profiling using tests from non-screening cognitive batteries, including the Free Recall Test (episodic memory), Simon Task (executive function), WAIS Block Design (visuospatial function), WMS Associate Learning, and the WAIS -R Digit Symbol Substitution Test. These domain-specific assessments enabled a rich, multimodal characterization of cognitive function associated with each mVC. For number of visits and follow up times see (Supplementary data: Neuropsychological data). The lab analysis methods for the CSF and plasma analysis are presented in detail elsewhere^5^. In summary, CSF Aβ42 and CSF phosphorylated tau (p-tau181) levels were measured with the fully automated Roche Elecsys immunoassay ^40,41^. Plasma Aβ42 and p-tau217 levels were measured on the Fujirebio Lumipulse G1200 automated immunoassay platform^42^. Plasma NfL and GFAP levels were measured on the SIMOA Quanterix HD-X high sensitivity immunoassay platform^43^.

### Simulation scheme and simulated data

To evaluate the robustness of the pipeline, we generated synthetic datasets with varying numbers of modalities, variables per modality, and individual, pairwise, and common mVCs (Batch 1, Figure S-4). Additionally, we tested increasing proportions of missing values, affecting either a single modality or all modalities (Batch 2, Figure S-5). Simulations featuring clustered observations across one or more mVCs demonstrated the pipeline’s ability to identify groups of subjects with similar multimodal patterns (Batch 3, Figure S-6). The data generation process generalizes the simulation algorithm used for three datasets by Gaynanova and colleagues^11^. Our evaluation is based on estimated rank performance compared to ground truth across the different batches. Specifically, for Batches 1 and 2, we calculated how many individual, pairwise, and common mVCs were identified by the model relative to the true ranks. For Batch 3, we computed the Pearson correlation between identified and ground truth mVCs to assess whether the SLIDE model accurately reconstructs clusters of observations under varying configurations. Simulation parameters not detailed here follow those in the original SLIDE implementation publication (positive scaling constants, signal-to-noise ratio, and standard uniformly distributed scores and loadings). All simulations were designed to reflect typical challenges encountered in biomarker discovery research. Specific simulation design parameters are summarized in the supplementary figures (Figure S-4/-5/-6).

### Multimodal pipeline overview in six steps

To effectively analyze the structural relationship between two or more biomarkers, observations in each biomarker modality should map to the other biomarker modalities in terms of an interpretable feature. Therefore, at a first step, the pipeline utilizes fixed and random effect linear modelling to align the different biomarker modalities (**step 1**). This is followed by missing data imputation (**step 2**), using the iterative singular value decomposition (SVD) algorithm by Fuentes and colleagues ^44,45^. This method was selected because SVD is well suited for high-dimensional data, has been applied in previous multiview analyses, and leverages the concatenated biomarker matrix rather than focusing solely on the modality with missing values^46^. Dataset standardization is essential, as variance decomposition using Structural learning and Integrative Decomposition (SLIDE) is not a scale-invariant process (**step 3**). It is carried out in two stages: within-modality and between-modality standardization. Within-modality standardization mitigates variable discrepancies within a modality, such as differences between MRI ROI volume (mm^3^) and cortical thickness (mm). We assume subject *i* ∈ {1, …, *n*}, modality *m* ∈ {1, …, *d*}, variable *j* ∈ {1, …, *p*_*m*_}. For variable *j* in modality m and dataset *X*_*m*_, 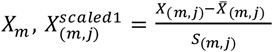, where 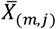 is the mean and *S*_*m,j*_ is the variable standard deviation. 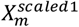 represents the concatenation of scaled and centered variables within dataset *X*_*m*_. Between modality standardization prevents scenarios where the largest modality dominates. After the between modality standardization, each modality *i* will be equal to 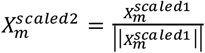, where ||*X*_*m*_ || is the Frobenius norm of *X*_*m*_. Each modality 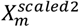 contributes equally to the total variation of the concatenated data. For simplicity, henceforth we refer to 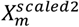 as *X*_*m*_. The aligned and standardized data are then fed into the SLIDE model ^32^ (**step 4**). SLIDE was selected as the backbone of the pipeline because it effectively disentangles sources of variance based on modality contributions. This capability is particularly valuable in biomarker discovery, as it allows researchers to pinpoint the origins of heterogeneity through biomarker variance. SLIDE generalizes classical low-rank factorization techniques such as Principal Component Analysis (PCA) and Joint and Individual Variation Explained (JIVE). This factorization approach explicitly models shared, partially shared, and modality-specific components within a unified framework. Following step 3, each *X*_*m*_ is linearly approximated by a set of latent vectors *U* (shared across subjects) and modality specific loading matrices *V*_*m*_, and an error matrix, representing residual noise:

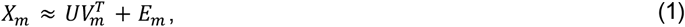

Unlike PCA and JIVE where the *V*_*m*_ matrix contains mixed loadings across all modalities, SLIDE introduces a structured sparsity pattern on the concatenated loading matrix *V* = [*V*_1_, …, *V*_*k*_ ], with *r* loading vectors, *r* ∈ {1, …, *k*}. *V* enables the model to capture not only global (latent factors shared across all modalities) and individual (latent factors with single-modality loadings) structures, but also partially shared structures (latent factors present in a subset of modalities). To formalize this generalization, equation (1) can be rewritten as

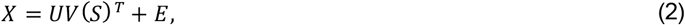

where *S* ∈ {0,1}^*dxr*^ is a binary “structure matrix” that encodes whether a modality participates in a latent component, while *V*(*S*) enforces the corresponding block-sparse pattern in *V*. An example of the S matrix for three modalities is shown below:

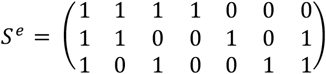

Here, the *S*^*e*^ matrix defines 7 multimodal variance components (mVCs). The first column represents a globally shared component of variance across all three modalities, while columns 2, 3, and 7 represent pairwise shared variance components. Finally, columns 4, 5, and 6 capture modality -specific variance components, representing variation unique to each individual modality.

An orthogonality constraint on *U* (*U*^*T*^ *U* = 1) ensures identifiability of scores and simplifies interpretation, as the resulting components are uncorrelated latent axes of variance. However, unlike PCA, the *V* matrix is not orthogonal, meaning the latent directions may not be mutually orthogonal. As a result, multiple latent components may explain overlapping sets of features, which can make interpretation more challenging. On the other hand, this non-orthogonality allows SLIDE to capture correlated or hierarchically structured latent sources, potentially reflecting true biological overlap. In this pipeline, expert supervision determines which latent components are selected for interpretation. SLIDE estimation involves three main step (see Gaynanova et al., 2019^11^ for details):

1. **Structure learning**: Matrix factorization with a sparsity-inducing penalty identifies candidate models by minimizing the following cost function

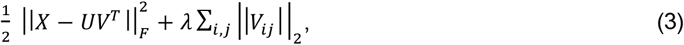

where *V*_*m,j*_ ∈ ℝ^*p*^ is the *j*_*th*_ column of the *m*-th block *V*_*m*_ ∈ ℝ^*p x r*^ of the loadings matrix *V*, with *r* = min (*n, p*). The parameter *λ* controls sparsity in loadings and determines which modalities contribute to each latent factor. In practice, we optimize across multiple *λ* values and select the best model using an approach similar to lasso variable selection (*L*_1_ regularization).
2. **Model fitting**: Given a structure S, alternating least squares is used to estimate *U* and *V* under the orthogonality constraint. This core step of SLIDE corresponds to a constrained low-rank approximation that jointly captures global, partially shared, and individual variation across multimodal data.
3. **Model selection**: Model fitting yields a set of candidate structures S. The optimal structure *S*^∗^ is chose via bi-cross-validation step, which evaluates reconstruction error across held-out data folds to ensure balanced model complexity and generalizability.

This decomposition produces interpretable latent factors (mVCs) that can be directly linked to biological processes shared across modalities, or unique to specific biomarker domains (axes of variance). Empirical studies show that SLIDE outperforms previous linked-component models in recovering ground-truth signals, estimating component ranks, and uncovering biologically meaningful structures in multi-omics datasets^32^. In our implementation, this framework was adapted to integrate neuroimaging and molecular biomarkers, facilitating the discovery of canonical multimodal Alzheimer’s disease factors (e.g., shared amyloid-β-tau-atrophy patterns) and secondary modality-specific trajectories that reflect cognitive, environmental, genetic, and regional heterogeneity.

In **step 5**, similar to PCA, and for the sake of parsimony, only a subset, not the entire mVC matrix will is interpreted. A singular value approximation, as used in PCA, would be applicable if SLIDE mVCs were not explicitly designed to reflect shared, pairwise, and individual modality structures. Therefore, we adopted an alternative approach that respects the structure of the SLIDE model. After estimating the latent score matrix *U* ∈ ℝ^*n x r*^ and loading matrix *V* ∈ ℝ^*p x r*^, we quantify the variance each mVC explains in the original data matrix *X* = [*X*_1_, …, *X*_*d*_ ]. For each component *U*_*r*_, *r* ∈ {1, …, *k*} and each variable *j* ∈ {1, …, *p*}, we model the original variable *x*_*j*_ as a linear function of the latent scores *u*_*r*_:

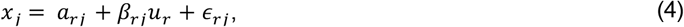

where *x*_*j*_ ∈ ℝ^*n*^ is a vector of variable *j* across *n* subjects, *u*_*r*_ ∈ ℝ^*n*^ is the score vector for mVC *r*, and *a*_*rj*_, *Β*_*rj*_ are the intercept and slope coefficients estimated by ordinary least squares (OLS). *ϵ*_*rj*_ is an error term. The coefficient of determination 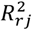 quantifies the proportion of variance in feature *x*_*j*_ explained by mVC *r*. The computation yields a matrix *R*^2^ ∈ ℝ^*p x r*^ describing the feature-level explanatory power of each mVC. To maintain consistency with the modality structure learned by SLIDE, only features with nonzero loadings in *V*_.*j*_ are considered active for component *j*:

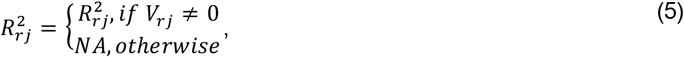

The resulting *R*^2^ matrix reflects the conditional variance explained within each mVC’s domain of influence. For mVC *r*,

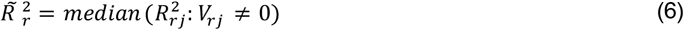

Median statistics were used instead of mean to provide a robust central measure of explanatory power, less affected by outliers and variable distributions. This statistic, henceforth called component’s variance proxy, reflects the typical variance explained per *R*^2^ feature by each mVC, enabling direct comparison of components’ relative contribution to the data structure *X*.

For the application to the neuroimaging datasets, an additional step (**step 6**) ensures reduced redundancy in the mVC interpretation. Since separate models were trained for cognitively impaired and unimpaired subjects, we removed highly correlated components and retained a single representative from each correlated group. This step operates on the subject-space score vectors (columns of the two *U* matrices, one for CU and one for CI) and applies a univariate Pearson correlation criterion, along with a scalar component score (e.g., the component’s variance proxy) to select representatives. To identify surviving components, we first computed the Pearson’s correlation coefficient matrix from the concatenated *U* matrices from both diagnostic groups. An mVC is considered unrelated other mVCs if its absolute Pearson correlation is below 0.4 with all other mVCs, and is therefore retained automatically (stage 1). Using the remaining mVCs, we determine which to retain using a greedy algorithmic approach. Starting from the first mVC in the matrix, we select only those with absolute correlation greater than 0.4. Within this candidate group, we retain the mVC that either explains the most multimodal information prioritizing global -> pairwise -> individual components or has the highest component variance (step 5). The remaining candidates are discarded, as they are correlated with retained mVCs and offer limited or no additional value. The process concludes when no further mVC candidates remain (stage 2). The final set of retained mVCs comprises the union of automatically independent components (Stage 1) and representatives selected from correlated groups (Stage 2).

#### Analysis flowchart

MRI, tau and amyloid-β-PET Freesurfer output datasets were aligned to a common timescale using linear modelling. Hierarchical models were used to estimate biomarker levels in the discovery set as follows

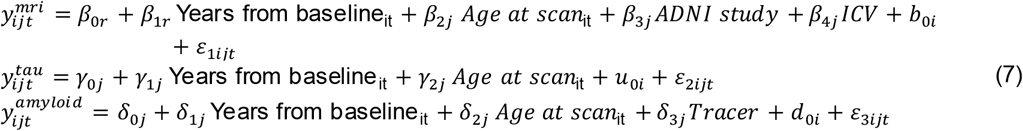

In equation (7), 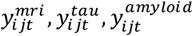 represent the MRI, tau, and amyloid-β measurements of subject *i*, in ROI *k*, at time *t*. Moreover, 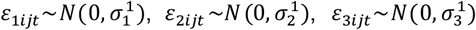 are the model residual distributions expressing population level error, and 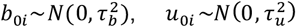, and 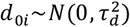 represent subject-specific variance distributions, reflecting individual-level deviations from 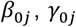, and 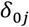 respectively. The other *Β, γ*, and *δ* parameters are population slopes. 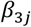 accounts for differences between ADNI studies (any ADNI1, 2, GO, 3 systematic Freesurfer differences) at a region of interest level, 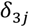 accounts for differences in intracranial volume (only in the 7 bilateral subcortical volumetric ROIs), and 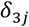 accounts for differences between the FBB and FBP tracers. Fixed and random effects were independently estimated using the standard linear mixed-effects optimization procedure (lmer function, lme4 package in R language). The models also included age at scan (at the first measurement) to account for biomarker differences associated with aging at the population level. The variable “years from baseline” refers to the difference between the age at scan and 75 years of age. Using these two temporal variables, we estimated the biomarker’s sample-level value for a specific ROI at age 75, with the slope representing the annual change beyond age 75. Lower in the model hierarchy, the 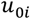 predictions return the individual’s specific deviation (best linear unbiased predictor) from the sample mean estimate at the age of 75 years. This model hierarchy, combined with longitudinal biomarker data, anchors all study participants to age 75 and provides a measure of individual variability relative to the overall sample. Following the statistical estimation in formula (7), all three biomarker datasets are expressed as random intercept variations, reducing the longitudinal dataset to a matrix ***X***: (*n, p*), where *n* is the number of subjects and *p* c is the number of ROIs (82 Desikan-Killiany and subcortical volumes). Specifically for OASIS, where tau PET data were only available cross -sectionally, formula (7) for 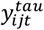 was adapted to a non-hierarchical model including only fixed effects, excluding 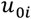 and 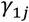. In OASIS, *u*_0*i*_ was replaced with estimated residuals *e*_2*ij*_ from the random error term *ε*_2*ij*_. These residuals resemble predicted random intercepts and capture subject-specific variation from 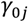 (The population average at age 75), albeit based on a single observation per subject.

Previous research in AD and healthy aging has shown that heterogeneity differs between CU and CI aging^15,33,47^. Based on the literature, we optimized three models in the discovery dataset: a diagnosis - agnostic model (*M*_*all*_), a CU only model (*M*_*CU*_), and a CI only model (*M*_*CI*_). We compared the results of the diagnosis-agnostic and diagnosis-specific models. The final results from the discovery dataset were validated using the OASIS dataset. However, since model predictions relied solely on pre-trained parameters from the discovery dataset, we also trained a separate de novo model (*M*_*V*_) on the validation set (diagnosis-agnostic only, due to the small CI sample) to explore multimodal biomarker heterogeneity in the validation cohort. The X data from each of the four models were processed through a six -step multimodal pipeline presented above, resulting in mVC scores and loading matrices *U*: {*U*_*all*_, *U*_*CU*_, *U*_*CI*_, *U*_*V*_}, and *V*: {*V*_*all*_, *V*_*CU*_, *V*_*CI*_, *V*_*V*_ }, respectively.

The model output matrices *U*, and *V* were used to visualize heterogeneous patterns in the discovery and validation datasets. For each mVC *k, U*_*k*_ represents a component summarizing **inter-subject** variability across participants. The models below assess how these latent factors relate to regional imaging values (subject-specific random intercepts from equation (7)) across diagnostic groups. The linear model used to visualize mVC *r*, for individual *i*, ROI *j*, modality *m* ∈ {*MRI, Amyloid, tau*}, and diagnosis group *g* is

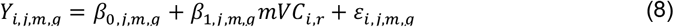

In equation (8), *Y*_*i,j,m,g*_ is the imaging signal (random intercept cortical thickness and subcortical volume, amyloid-β/tau SUVR) for subject *i* in region *j* and modality *m*, restricted to group *g, mVC*_*i,r*_ is the *U*_*i,r*_ score of individual *i* in component *r, Β*_1,*j,m,g*_ is the maximum likelihood slope estimate representing the association between *U*_*i,r*_ scores and regional imaging measure, and *ε*_*i,j,m,g*_∼*N*(0, *σ* ^2^), is the residual associated to *Y*_*i,j,m,g*_. Importantly, the primary parameter of interest *Β*_1,*j,m,g*_, the effect size of the latent factor on the imaging signal was visualized. A two tailed t -test was used to assess *H*_0_: *Β*_1,*j,m,g*_ = 0 and the resulting p-values were corrected for multiple comparisons across all within-modality ROIs using the Benjamini–Hochberg FDR procedure with *q* = 0.05. At a confidence level of *a* = 0.05, ROIs with *p*_*j* ,*m,g*_ < *a* were retained for visualization, others were set to zero (visualized as gray color). For retained regions (mVCs were unit-scaled and centered), the difference of fitted values between - 2 SD and +2 SD from the mean were visualized to illustrate individual differences as *U*_*i,r*_ scores increase. To facilitate comparison across mVCs, all *U*_*i,r*_ scores were ordered such that lower values correspond to lower *AΒ* levels. The *Β* coefficients were directly mapped to color intensity, reflecting both the direction and magnitude of the association. *Β* values were truncated using fixed thresholds based on the random intercept range of the CU dataset (from formula 7) to preserve image contrast (see color legends in Fig 1A/2A). ROIs with large absolute difference between -2 and +2 SD from the mean indicate strong associations with the mVC. This visualization reveals whether a given mVC component reflects large variations in structural amyloid-β deposition, tau accumulation, or atrophy (structural anatomy). The R packages ggplot2, ggseg, and dplyr were used for visualization.

#### Power analysis

For the application in the discovery dataset (N = 829 rows and p = 246 variables, a 3.36 N:p ratio) data points exceeded the number of variables in each separate modality (10.1 subjects per single modality). Given that the output model included 11 factors, the variable-to-factor ratio was 22.3. Research in latent factor analysis proposes that a rule of thumb of 300 samples is deemed a sufficient sample, while 1000 samples is considered excellent ^48^. Moreover, a variable-to-factor ratio of at least 7 is considered a safe minimum sample size recommendation for conducting factor analyses ^49,50^. To be precise, we chose to simulate datasets with 300 observations (See Fig. S-3 to S-6, Tables S-10/-11) to test whether mVC could be accurately identified. The simulated results showed that the pipeline only failed to identify the correct mVC when more than seven modalities were utilized, which is not the case with ADNI or OASIS were three modalities where used. Therefore, the number of observations (1250 individuals and 246 variables in total) is deemed more than sufficient to discover meaningful multimodal associations using the proposed pipeline in this application.

#### Characterization of Latent Factors Across Genetic, Environmental, Fluid (CSF and Plasma) Biomarkers, and Cognitive Domains

We characterized the discovered mVCs in relation to key demographic, environmental, and genetic factors including biological sex, APOE genotype, and education level. Following the same statistical framework as the imaging analyses, we tested the association of these factors with increasing *U*_*r*_ scores. Accounting for education level and age at cognitive assessment (anchored to age 75) we modelled cognitive performance using the following specification:

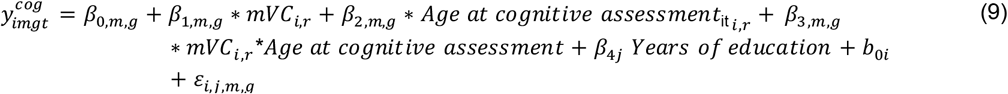

In formula (9), the link function was either Gaussian or Poisson-distributed depending on the response variable’s distribution (lmer and glmer function, lme4 package in R language). The primary parameters here of interest *Β*_1,*m,g*_ and *Β*_3,*m,g*_, capture the change in cognitive performance associated with a one - unit increase in mVC score, and the interaction between mVC and age, respectively. The other parameters are similar to formula (7). P-values were adjusted across mVCs using the same procedure as in formula (8). Regarding the CSF/plasma/genetic markers and following formula (8), we employed either a Gaussian or Bernouli-distribution for linear (Tau PET meta-temporal SUVR, Amyloid-β-PET centiloid, CSF p-tau181/Aβ42, plasma p-tau217/Aβ42, NFl, GFAP) and dichotomous (APOE e4 homo/hetero-zygosity) outcomes respectively. In all cases we assessed whether a shift from -2 to +2 SD in mVC scores corresponds to a significant change in each outcome, adjusting for age at sampling and correcting for multiple comparisons. For plasma, CSF and APOE outcomes a single observation for each subject was used, a significant result means that by shifting from subjects on the lower towards subjects on the higher mVC scores’ end, the marker changes significantly.

#### Latent Factors in Relation to Disease Stage and Progression

To examine how mVC components relate to diagnostic progression over time, we categorized participants based on their clinical trajectory between their first and last available diagnosis. Specifically, we identified individuals who remained cognitively unimpaired (CU to CU), as well as those who progressed from CU to mild cognitive impairment (CU to MCI) or dementia due to Alzheimer’s disease (CU to AD). Additionally, we included individuals who transitioned from MCI to CU or AD, and those who remained stable (MCI to MCI). For each participant, a single observation was retained, and analyses focused on the three primary CU - based transition groups (Supplementary data: Diagnosis progression). For each mVC component, we fitted separate multinomial models (similar to formula 8) using diagnostic change as the dependent variable and the component score as the independent variable. The models were fitted with 5-fold cross validation to account for differences sample sizes in cognitive progression (transition) (multinom function, nnet package in R language). The two models included CU to CU or MCI to MCI as the reference group, depending on the transition set. P-values were adjusted across components using the Benjamini–Hochberg procedure, consistent with the approach described in formula (8). To preclude circularity/data leakage, we enforced the following: (1) unsupervised estimation of the mVCs was computed from imaging data alone, without diagnostic labels or transition outcomes inside the model. (2) predictors were neuroimaging baseline scores (at the age of 75), while outcomes were future transitions. Similarly with the fluid markers assessed with Bernoulli-distributed outcomes, we report significant change in the odds ratio for shifting from a stable diagnosis to regression or progression for a shift from -2 to +2 SD in mVC scores.

All statistical analyses were conducted in R (version 4.5.0). All experiments described in the methods section were conducted on a Linux Mint system equipped with a single 8-core CPU (Intel i7-7829HQ, 2.9 GHz).

## Data Availability

see Data availability section

## Data availability

All derived mVC scores used in this study are indexed by participant ID are available in the supplementary material. The raw imaging and clinical data were obtained from the ADNI and OASIS datasets under data-sharing agreements. These agreements do not permit redistribution of the original data. Researchers interested in accessing the full datasets must apply directly to the respective data providers. Data from the Alzheimer’s Disease Neuroimaging Initiative (ADNI) are publicly available at https://adni.loni.usc.edu upon registration and agreement to the data use terms. Data from the OASIS- 3 study can be requested via https://www.oasis-brains.org/. Once access is granted, investigators can match our derived scores to the corresponding subject IDs in ADNI and OASIS. For assistance with matching (see supplementary data; subject specific mVC scores and mVC loadings per dataset) or implementation, researchers may contact the corresponding author, K.P. Additionally, the full data processing pipeline and model application steps are described in the Methods section, and an example script is provided in the associated GitHub repository to enable replication of the mVC derivation on other independent datasets.

## Ethics statement

All data used in this study were obtained from the Alzheimer’s Disease Neuroimaging Initiative (ADNI) and the Open Access Series of Imaging Studies (OASIS-3). For both cohorts, study protocols were approved by the appropriate Institutional Review Boards or ethics committees at the participating institutions, and all participants or their legally authorized representatives provided written informed consent prior to study enrollment, in accordance with applicable institutional, national, and federal guidelines. Data were shared for secondary analysis in de-identified form only, under formal Data Use Agreements governing ethical use, confidentiality, and data protection, and prohibiting participant re-identification.

## Acknowledgments

Data used in the preparation of this article were obtained from the Alzheimer’s Disease Neuroimaging Initiative (ADNI) database (https://adni.loni.usc.edu)^51^. The investigators within ADNI contributed to the design and implementation of ADNI and/or provided data but did not participate in the analysis or writing of this report. A complete listing of ADNI investigators can be found at: https://adni.loni.usc.edu/wp-content/uploads/how_to_apply/ADNI_Acknowledgement_List.pdf.

Data were also provided in part by OASIS-3^37^: Longitudinal Multimodal Neuroimaging (Open Access Series of Imaging Studies). OASIS-3 data were collected and shared by Washington University in St. Louis under the leadership of principal investigators Tammie L.S. Benzinger, Daniel Marcus, and John C. Morris. OASIS-3 is supported by the National Institutes of Health through grants P50 AG00561, P30 NS098577, P01 AG026276, P01 AG003991, R01 AG043434, UL1 TR000448, and R01 EB009352. Florbetapir F18 (AV-45) and Flortaucipir F18 (AV-1451) PET tracer doses were provided by Avid Radiopharmaceuticals, a wholly owned subsidiary of Eli Lilly and Company.

The following funding bodies have contributed to this project: K.P has received funding from the Swedish Research Council, Gamla Tjänarinnor Foundation, Erik and Edith Fernström’s Foundation, Stohnes Foundation, Foundation for Geriatric diseases at Karolinska Institute, Loo and Hans Osterman Foundation, Strategic Research Program in Neuroscience Foundation.

## Notes

**Competing Interest Statement:** We declare no conflicts of interest.

### Competing Interest Statement

The authors have declared no competing interest.

### Author Declarations

Ethical committees (IRBs) have approved the collection of data for the ADNI and OASIS public datasets utilized in this study (see Ethical statement). Local ethical approvals were received for this study.

