## Supplementary tables and figures for "Multimodal axes reveal individualized amyloid-β, tau, and neurodegeneration coupling in aging and Alzheimer’s disease"

* Corresponding author: Konstantinos Poulakis

**Competing Interest Statement:** We declare no conflicts of interest.

**Classification:** **Major**, Biological Sciences; **Minor,** Neuroscience.

**Keywords:** Alzheimer's disease, unsupervised learning, multi-view analysis, amyloid-β-PET, tau PET, MRI

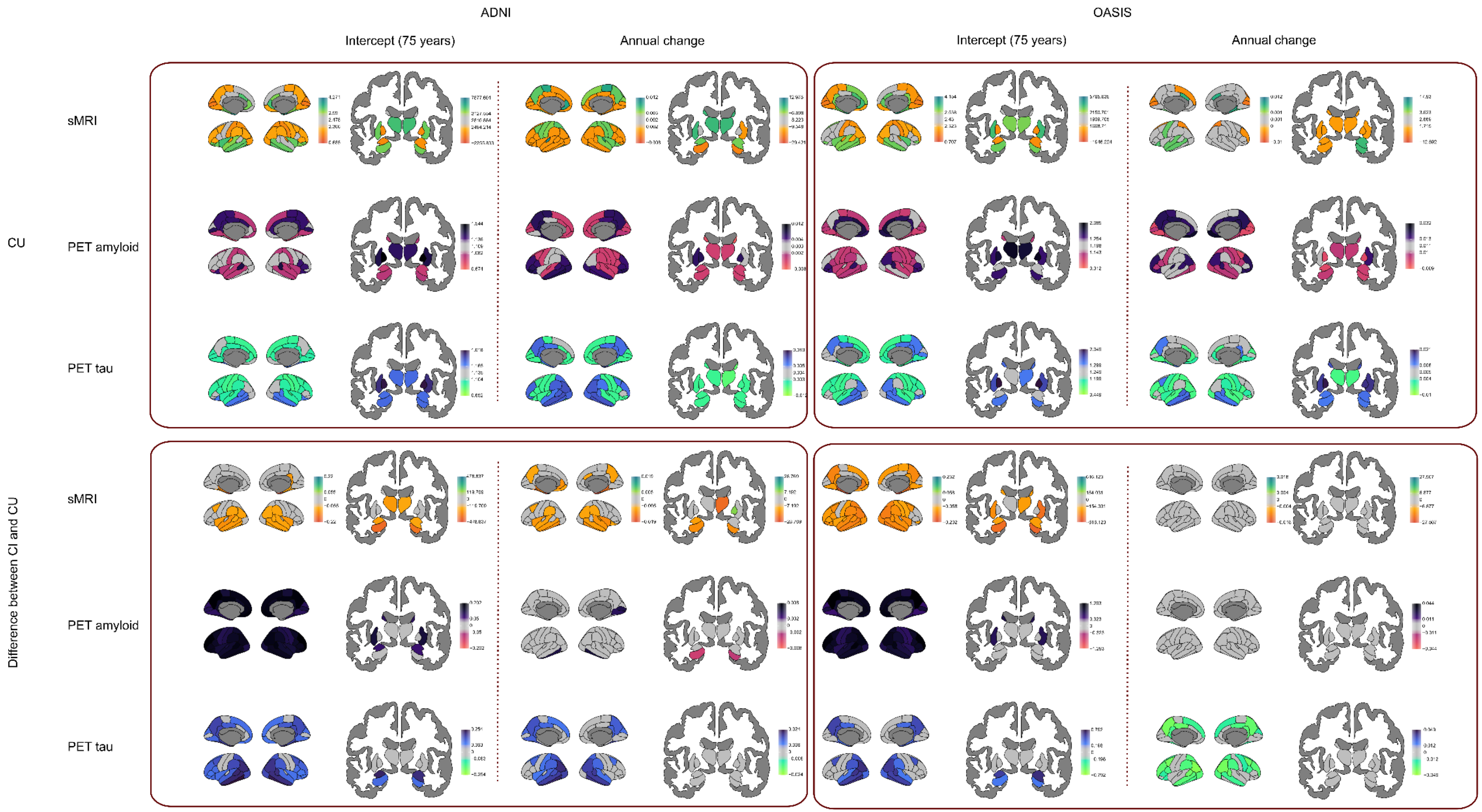


**Figure S-1. Population estimates of expected structural MRI, Amyloid-β and Tau PET intercepts and slopes over time**. Brain maps that capture between dataset and clinical stages patterns of cortical thickness/volume (T1 MRI), amyloid-β PET (SUVR), and tau PET (SUVR). ROI estiamtes in the space of each biomarker are FDR-corrected (Benjamini–Hochberg, q<0.05).

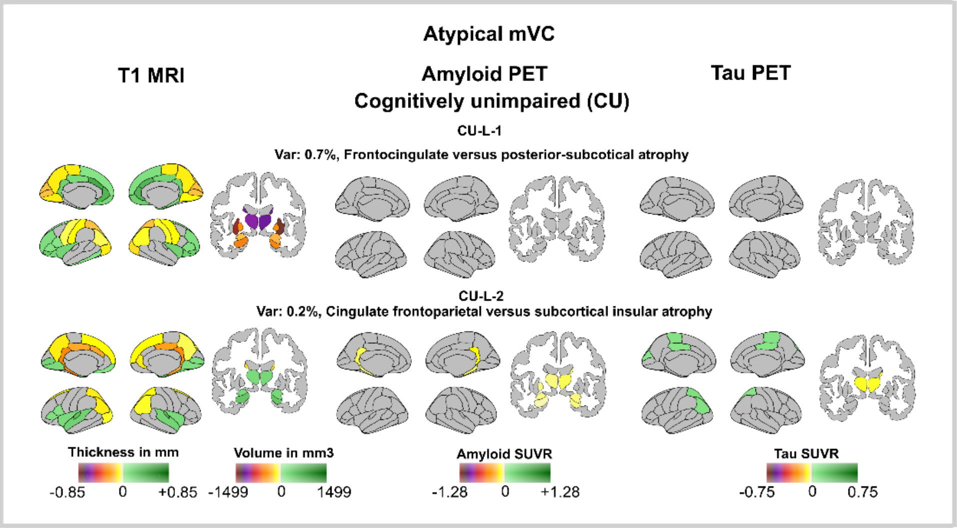


**Figure S-2. Population estimates of expected structural MRI, Amyloid-β and Tau PET intercepts and slopes over time**. Shown are brain maps of multimodal variance components (mVCs) that capture the low variance axes of A-T-N(atrophy) coupling in cognitively unimpaired (CU; CU-L-1, CU-L-2) participants. For each axis, cortical thickness/volume (T1 MRI), amyloid-β PET (SUVR), and tau PET (SUVR) are visualized as ROI wise contrasts between individuals at −2 vs +2 SD on the mVC score (Methods, Eq. 8). non significant ROIs are gray. Color bars reflect fixed truncation based on the CU random intercept ranges to preserve contrast across mVCs. Axes were estimated in stage specific multiview models (separate CU and CI fits) following longitudinal mixed effects harmonization (age anchored at 75 y; intercepts used for visualization) and SLIDE decomposition; the variance proxy noted per axis summarizes relative contribution to multimodal structure. Baseline CU and CI A-T-N(atrophy) profiles are shown in Fig. S-1).

| | CU | CI | CI : MCI | CI : AD |
|---|---|---|---|---|
| N, N(%) | 477 (57.5%) | 352 (42.5%) | 264 (75%) | 88 (25%) |
| Females, N(%) | 278 (58.3%) | 146 (41.5%) | 110 (41.7%) | 36 (40.9%) |
| Age, median(mad) | 75 (8.5) | 76.6 (7.9) | 77.4 (7.9) | 74.6 (7.9) |
| Years of education, median(mad) | 16.65 (2.37) | 16.15 (2.6) | 16.24 (2.55) | 15.86 (2.71) |
| Apoe e4 allele carrier, N(%) | 157 (32.9%) | 159 (45.2%) | 107 (40.5%) | 52 (59.1%) |
| Apoe e2 allele carrier, N(%) | 63 (13.2%) | 33 (9.4%) | 25 (9.5%) | 8 (9.1%) |
| MMSE | 28.9 (0.05) | 26.2 (-0.08) | 27.4 | 22.6 |
| CDR Sum Of Boxes | 0.27 (0) | 1.89 (0.081) | 1.35 | 5.49 |
| **ADSP** | | | | |
| Memory function | 0.98 (-0.031) | 0 (-0.035) | 0.27 | -0.73 |
| Executive function | 0.77 (-0.021) | 0.17 (-0.029) | 0.34 | -0.33 |
| Language function | 0.78 (-0.024) | 0.26 (-0.032) | 0.4 | -0.19 |
| Visuospatial function | 0.57 (-0.008) | 0.26 (-0.013) | 0.46 | 0 |
| **MoCA** | | | | |
| Memory | 2.1 (-0.06) | 0.5 (-0.01) | 0.7 | 0.1 |
| Attention | 4.9 (0) | 4.6 (0) | 4.7 | 4.2 |
| Language | 4.6 (-0.02) | 4.2 (-0.01) | 4.3 | 3.8 |
| Executive | 3.6 (-0.01) | 3 (0) | 3.1 | 2.5 |
| Visuospatial | 3.5 (0) | 3.1 (-0.02) | 3.2 | 2.5 |

**Table S-1.Supplementary table. Main dataset (ADNI) characteristics.** All cognitive estimates are projected at the age of 75 years, with annual slopes in parentheses. For MoCA and ADSP sub-domains, Poisson and linear (Gaussian) mixed effects regressions were used. Only significant baseline and slope estimates are reported (0.05 significance level (see methods). MMSE: mini mental state examination; CDR: clinical dementia rating scale; ADSP cognition: z values (lower is worse); MoCA, higher equals better scoring, memory: delayed recall (0-5), Attention: digit span forward/backward, tap letter A, serial 7s subtraction (0-6); Language: naming, repetition (0-5), Executive: trail making, verbal fluence (0-4); Visuospatial: cube copy, clock drawing (0-4).

| | CU | CI |
|---|---|---|
| N, N(%) | 363 (86.2%) | 58 (13.8%) |
| Females, N(%) | 214 (59%) | 28 (48.3%) |
| Age, median(mad) | 65.8 (8.4) | 71.2 (7.5) |
| Years of education, median(mad) | 16.53 (2.26) | 15.07 (2.77) |
| Apoe e4 allele carrier, N(%) | 130 (35.8%) | 38 (65.5%) |
| Apoe e2 allele carrier, N(%) | 57 (15.7%) | 4 (6.9%) |
| MMSE | 29.12 (0) | 26.02 (0) |
| CDR Sum Of Boxes | 0.0675 (0) | 3.375 (0.525) |
| Recall test freely (episodic) | 29.63 (-0.16) | 0 (-0.65) |
| Simon task (executive) | 97.6 (0) | 79.04 (0.293) |
| **MoCA** | | |
| Memory | 3.23 (-0.034) | 0.6 (0) |
| Attention | 5.64 (0) | 5.09 (0) |
| Language | 4.51 (0) | 4.27 (0) |
| Executive | 3.32 (0) | 2.7 (0) |
| Visuospatial | 3.48 (0) | 2.96 (0) |

**Table S-2. Supplementary table. Validation dataset (OASIS) characteristics.** All cognitive estimates are projected at the age of 75 years, with annual slopes in parentheses. Only significant baseline and slope estimates are reported (0.05 significance level (see methods). MMSE (for MMSE, Poisson mixed effects regression was used): mini mental state examination; CDR (for CDR, logistic mixed effects regression was used): clinical dementia rating scale; MoCA (for MoCA sub-domains, Poisson mixed effects regressions were used), higher equals better scoring, memory: delayed recall (0-5), Attention: digit span forward/backward, tap letter A, serial 7s subtraction (0-6); Language: naming, repetition (0-5), Executive: trail making, verbal fluence (0-4); Visuospatial: cube copy, clock drawing (0-4).

| | MMSE[2] | ADSP[1] | | | | MoCA[2] | | | | |
|---|---|---|---|---|---|---|---|---|---|---|
| mVC | | Memory | Language | Executive | Visuospatial | Memory | Language | Executive | Visuospatial | Attention |
| **Typical CU** | | | | | | | | | | |
| CU-T-1 | - | 0(-0.02) | 0(-0.01) | 0(-0.01) | - | 0(-0.11) | - | - | - | - |
| CU-T-2 | - | 0(-0.03) | 0(-0.02) | 0(-0.01) | -0.34(-) | 0(-0.09) | - | - | - | - |
| **Typical CI** | | | | | | | | | | |
| CI-T-1 | -5.58(-) | -1.62(-0.03) | -0.8(-0.01) | -1.16(-0.01) | -0.56(-) | -1.03(-0.02) | -0.61(-) | -0.42(-) | -0.81(-) | -0.58(-) |
| CI-T-2 | -4.02(-) | -0.99(-0.02) | -0.37(-0.01) | -0.75(-0.01) | -0.36(-) | -0.75(-0.02) | - | - | -0.61(-) | - |
| CI-T-3 | -4.43(-) | -1.18(-0.02) | -0.44(-0.01) | -0.81(-0.01) | -0.35(-) | -0.91(-0.02) | - | - | -0.62(-) | - |
| **Atypical CU** | | | | | | | | | | |
| CU-A-1 | - | - | - | - | - | - | - | - | - | - |
| **Atypical CI** | | | | | | | | | | |
| CI-A-1 | -3.32(-) | -0.81(-0.01) | -0.51(-0.01) | -0.43(-0.01) | -0.25(-) | -0.62(-) | -0.66(-) | -0.44(-) | -0.51(-) | - |
| CI-A-2 | -4.81(-0.16) | -1.25(-0.03) | -0.67(-0.01) | -0.41(-0.02) | 0(-0.01) | -1.07(-0.02) | -0.62(-) | -0.51(-) | -0.45(-0.03) | - |
| CI-A-3 | -1.69(-) | -0.35(-0.01) | -0.24(-) | -0.28(-) | - | - | - | - | - | - |
| **Low variance components** | | | | | | | | | | |
| CU-L-1 | - | - | - | 0(0.01) | 0(0.01) | - | - | - | - | - |
| CU-L-2 | - | 0(0.01) | 0.25(0.01) | - | - | - | - | - | - | - |

**Table S-3. Longitudinal association between the ADNI Montreal Cognitive assessment scale (MoCA)/ADSP cognitive scores battery and the multimodal variance component scores (mVC scores)**. Estimated difference in cognition between -2 and +2 standard deviations in mVC scores. Only significant mean differences (longitudinal annual cognitive decline for the same shift in mVC scores) after FDR correction at 0.05 significance level are presented (see methods).

[1]Linear mixed effects regressions were used for estimation.

[2]Poisson mixed effects regressions were used for estimation.

| mVC | Word Recall | Following Commands | Constructional Praxis | Delayed Word Recall | Naming Objects and Fingers | Ideational Praxis | Orientation | Word Recognition | Remembering Test Instructions | Comprehension of Spoken Language | Word finding Difficulty | Language | Number Cancellation |
|---|---|---|---|---|---|---|---|---|---|---|---|---|---|
| **Typical CU** | | | | | | | | | | | | | |
| CU-T-1 | 0(0.3) | - | - | 0(0.1) | - | - | 0(0.01) | - | - | - | - | - | - |
| CU-T-2 | 0(0.3) | - | - | 0(0.12) | - | - | - | 0(0.1) | - | - | - | - | - |
| **Typical CI** | | | | | | | | | | | | | |
| CI-T-1 | 7.8(0.3) | 0.2(-) | 0.4(-) | 4.1(-) | 0.1(-) | 0.2(-) | 1.2(0.1) | 3.6(0.1) | 0.1(0.01) | 0.11(-) | 0.3(-) | 0.04(-) | - |
| CI-T-2 | 5.8(-) | 0.1(-) | 0.3(-) | 3.4(-) | - | 0.1(-) | 0.8(-) | 2.3(-) | 0.1(-) | 0.1(-) | 0.3(-) | 0.02(-) | - |
| CI-T-3 | 6.7(-) | 0.1(-) | - | 4(-) | - | 0.1(-) | 0.9(-) | 2.9(-) | 0.1(-) | 0.1(-) | 0.3(-) | 0.02(-) | 0.8(-) |
| **Atypical CU** | | | | | | | | | | | | | |
| CU-A-1 | - | - | - | - | - | - | - | - | - | - | - | - | - |
| **Atypical CI** | | | | | | | | | | | | | |
| CI-A-1 | 4.3(-) | 0.2(-) | 0.3(-) | 3(-) | 0.1(-) | 0.1(-) | 0.4(0.1) | 2.2(-) | 0.04(-) | 0.1(-) | 0.2(-) | - | 0.4(-) |
| CI-A-2 | 5.9(0.3) | 0(0.01) | - | 3.6(0.2) | 0.2(0.01) | 0.1(0.01) | 0.9(0.1) | 3.8(0.1) | 0.04(-) | 0.1(-) | 0.2(0.02) | 0.03(-) | 0(0.1) |
| CI-A-3 | 2.2(-) | - | - | 1.1(-) | - | - | 0.3(-) | - | - | - | - | - | - |
| **Low variance components** | | | | | | | | | | | | | |
| CU-L-2 | 0(0.1) | - | - | - | - | - | - | - | - | - | - | - | - |

**Table S-4**. **Longitudinal association between the ADNI ADAS cognitive scores battery and the multimodal variance component scores (mVC scores)**. Estimated difference in cognition between -2 and +2 standard deviations in mVC scores. Only significant mean differences (longitudinal annual cognitive decline for the same shift in mVC scores) after FDR correction at 0.05 significance level are presented (see methods).

| mVC | APOE e4[1] | APOE e4 e4[1] | Years of education[2] | Tau PET meta-temporal SUVR [2] | Amyloid-β PET centiloid[2] | Plasma p-tau217/Aβ42[2] | CSF p-tau181/Aβ42[2] | Plasma NfL[2] | Plasma GFAP[2] |
|---|---|---|---|---|---|---|---|---|---|
| **Typical CU** | | | | | | | | | |
| CU-T-1 | - | - | - | 0.59(0) | 60.74(2.8) | 0.01(0) | 0.02(0) | - | - |
| CU-T-2 | - | - | - | 0.22(0) | 59.36(2.8) | - | 0.02(0) | - | - |
| **Typical CI** | | | | | | | | | |
| CI-T-1 | 0.02(1.1) | 0.06(1.2) | - | 0.93(0) | 156.15(1) | 0.04(0) | 0.08(0) | 6.67(0.6) | 166.84(6.6) |
| CI-T-2 | 0.03(1.1) | 0.07(1.1) | - | 0.66(0) | 160.8(0.4) | 0.03(0) | 0.06(0) | - | 126.26(6.6) |
| CI-T-3 | 0.02(1.1) | 0.04(1.2) | - | 0.68(0) | 161.47(0.5) | 0.03(0) | 0.07(0) | - | 146.67(6.7) |
| **Atypical CU** | | | | | | | | | |
| CU-A-1 | - | - | - | - | - | - | -0.01(0) | - | - |
| **Atypical CI** | | | | | | | | | |
| CI-A-1 | - | - | - | 0.36(0) | 24.21(2.3) | 0.01(0) | 0.03(0) | - | 77.21(6.9) |
| CI-A-2 | 0.34(1.1) | - | - | 0.42(0) | 27.45(2) | 0.01(0) | 0.03(0) | 10.63(0.6) | 89.81(6.4) |
| CI-A-3 | - | - | - | 0.29(0) | - | 0.01(0) | - | - | - |
| **Low variance components** | | | | | | | | | |
| CU-L-1 | - | - | - | 0.04(0.0) | - | - | - | - | 52.6(4.4) |
| CU-L-2 | 0.2(1.16) | - | - | -0.07(0) | - | - | - | - | - |

**Table S-5. Association between the ADNI genetic, environmental, CSF, and plasma markers with multimodal variance component scores (mVC scores)**. Estimated cross-sectional differences in various measures between subjects with -2 and +2 standard deviations in mVC scores. Only significant mean differences (standard error) after FDR correction at 0.05 significance level are presented (see methods).

[1]A logistic link was used, that is, the lower the value is, the higher the chances of being positive increase with increased mVC scores.

[2]A linear link was used, that is, the higher the table value is the more the response increases with increased mVC scores.

| mVC | Tau PET meta-temporal SUVR | Amyloid-β PET centiloid |
|---|---|---|
| **Typical CU** | | |
| CU-T-1 | 0.94(0.59) | -9.24(60.74) |
| CU-T-2 | 1.1(0.22) | -10.83(59.36) |
| CI-T-1 | 0.84(0.93) | -47.21(156.15) |
| CI-T-2 | 1.01(0.66) | -48.68(160.8) |
| CI-T-3 | 0.99(0.68) | -50.02(161.47) |
| **Atypical CU** | | |
| CU-A-1 | - | - |
| **Atypical CI** | | |
| CI-A-1 | 1.19(0.36) | 32.1(24.21) |
| CI-A-2 | 1.14(0.42) | 29.18(27.45) |
| CI-A-3 | 1.25(0.29) | - |
| **Low variance components** | | |
| CU-L-1 | - | - |
| CU-L-2 | 1.25(-0.07) | - |

**Table S-6. Tau and amyloid-β at low and high ends of the multimodal variance components (mVC).** Estimated markers at subject mVC score equal to -2 standard deviations from mVC center (difference between -2 and +2 SD). The estimates are based on linear regression with Gaussian link accounting for age at the image acquisition.

| mVC | CU to MCI versus CU to CU | CU to AD versus CU to CU | MCI to CU versus MCI to MCI | MCI to AD versus MCI to MCI |
|---|---|---|---|---|
| **Typical CU** | | | | |
| CU-T-1 | 0.159 | 0.002 | 17.308 | 0.044 |
| CU-T-2 | 0.035 | 0.001 | 80.531 | 0.062 |
| CI-T-1 | 0.065 | 0.001 | 44.484 | 0.029 |
| CI-T-2 | 0.206 | 0.043 | 12.62 | 0.078 |
| CI-T-3 | 0.135 | 0.02 | 12.419 | 0.073 |
| **Atypical CU** | | | | |
| CU-A-1 | 0.225 | 3656.851 | - | - |
| **Atypical CI** | | | | |
| CI-A-1 | 0.165 | 0.015 | 0.998 | 0.096 |
| CI-A-2 | 0.309 | 0.00 | 8.981 | 0.161 |
| CI-A-3 | - | - | - | - |
| **Low variance components** | | | | |
| CU-L-1 | - | - | - | - |
| CU-L-2 | 25.119 | 8.594 | - | - |

**Table S-7. Multimodal variance component (mVC) scores relative to diagnosis progression.** This table presents cross-validated odds ratios (ORs) estimating the likelihood of diagnostic transitions as multivariate component (mVC) scores increase from −2 to +2 standard deviations, derived from a 5-fold cross-validated multinomial logistic regression with FDR-corrected p-values. ORs below 1 indicate that higher mVC scores are associated with a greater probability of progressing to a more impaired diagnostic category, whereas ORs above 1 indicate a tendency to remain in the same diagnostic stage rather than progress or regress. For CU individuals, components with the lowest ORs are those most strongly associated with CU→MCI or CU→AD progression. CU-T-2 (reflecting widespread tau deposition) shows the strongest link to CU→MCI, followed by CI-T-1, while CU-L-2 shows ORs above 1, indicating a pattern more associated with remaining CU rather than progressing. Regarding CU→AD, ORs are generally lower than for CU→MCI, reflecting that several components track more advanced disease severity. Very high ORs for components like CU-A-1 suggest patterns unlikely to represent AD-type progression, whereas very low ORs for CI-A-2 highlight temporo-tau patterns closely linked to CU→AD transition. For MCI→CU, most ORs exceed 1, indicating that increasing mVC scores correspond to lower likelihood of clinical improvement and a greater chance of remaining stable MCI. In contrast, ORs for MCI→AD tend to fall below 1, consistent with the interpretation that increasing mVC expression, often reflecting structural or tau and/or amyloid-related pathology, is associated with a higher likelihood of progression to AD. Together, these cross-validated ORs summarize how each mVC relates to diagnostic stability or decline. Lower ORs identify mVCs linked to clinical worsening, whereas higher ORs point to patterns associated with stability or unlikely disease trajectories.

| | MoCA | | | | | | | | | |
|---|---|---|---|---|---|---|---|---|---|---|
| mVC | Memory | | Language | | Executive | | Visuospatial | | Attention | |
| **Typical CU** | CU | CI | CU | CI | CU | CI | CU | CI | CU | CI |
| CU-T-1 | - | -0.54(-) | - | - | - | - | - | - | - | - |
| CU-T-2 | - | -0.54(-) | - | - | - | - | - | - | - | - |
| **Typical CI** | | | | | | | | | | |
| CI-T-1 | -0.36(-) | -0.63(-) | - | - | - | - | - | - | - | - |
| CI-T-2 | -0.26(-) | -0.56(-) | - | - | - | - | - | - | - | - |
| CI-T-3 | -0.32(-) | -0.58(-) | - | - | - | - | - | - | - | - |
| **Atypical CU** | | | | | | | | | | |
| CU-A-1 | - | - | - | - | - | - | - | - | - | - |
| **Atypical CI** | | - | | | | | | | | |
| CI-A-1 | - | | - | - | - | - | - | - | - | - |
| CI-A-2 | -0.31(-0.06) | - | - | - | - | - | - | - | - | - |
| CI-A-3 | - | - | - | - | - | - | - | - | - | - |
| **Low variance components** | | - | | | | | | | | |
| CU-L-1 | - | | - | - | - | - | - | - | - | - |
| CU-L-2 | - | - | - | - | - | - | - | - | - | - |

**Table S-8**. **Longitudinal association between the Montreal cognitive assessment (MoCA) battery applied to OASIS participants and the multimodal variance component scores (mVC scores) extracted from the ADNI analysis**. Estimated difference in cognition between -2 and +2 standard deviations in mVC scores. Only significant mean differences (longitudinal annual cognitive decline for the same shift in mVC scores) after FDR correction at 0.05 significance level are presented (see methods). Poisson mixed effects regressions were used for estimation.

| mVC | MoCA Memory | | WAIS Block Design | | WAIS III Similarities | | WMS Associate Learning (summary score) | | WAIS-R Digit Symbol | | Simon task | | Recall tested freely | | Benson Drawings total score for copy | | Benson Drawings total score for delay | |
|---|---|---|---|---|---|---|---|---|---|---|---|---|---|---|---|---|---|---|
| | CU | CI | CI | CU | CU | CI | CU | CI | CU | CI | CU | CI | CU | CI | CU | CI | CU | CI |
| MRI mVC 1 | - | - | - | -3.46 | 0.97(0.16) | - | - | - | - | -5.64 | - | - | - | - | - | - | 0(-0.02) | - |
| MRI and Tau mVC | - | - | - | -3.45(-0.06) | - | - | - | - | -1.56 | -3.08(-0.15) | - | - | - | - | - | -0.91 | - | - |
| MRI and Amyloid and Tau mVC | -0.36 | -1.02 | - | -6.33 | - | - | 0(-0.16) | -2.22 | 0(-0.6) | -5.35 | - | - | 0(-0.33) | -4.25(-0.51) | - | -0.76 | -0.93(-0.16) | -1.9 |
| MRI mVC 2 | - | -0.5 | -1.03(-) | -4.66 | - | - | - | - | -1.96(-) | -4.3 | - | - | - | -3.78 | - | -0.95 | - | -1.09 |
| MRI mVC 3 | - | -0.56 | - | 0(-0.87) | -0.87 | - | 0(-0.05) | - | - | 0(-1.09) | - | - | - | -3.37 | - | 0(-0.12) | 0(-0.12) | -1.39 |
| Amyloid mVC 1 | -0.26 | -0.56 | - | -5.48 | 0(0.18) | - | 0(-0.12) | - | - | -3.79 | - | - | 0(-0.27) | -3.88(-0.55) | - | - | -0.79(-0.15) | -1.42 |
| Amyloid mVC 2 | -0.27 | -0.58 | - | -5.42 | 0(0.16) | - | 0(-0.14) | -1.95 | - | -3.96 | - | - | 0(-0.28) | -4.1(-0.54) | - | - | -0.77(-0.16) | -1.48 |
| Amyloid mVC 3 | -0.34 | -0.58 | - | -4.96 | - | - | 0(-0.14) | - | 0(-0.58) | -3.29 | - | - | -0.79(-0.29) | -3.87(-0.55) | - | - | -0.86(-0.16) | -1.42(-0.1) |
| Tau mVC 1 | -0.23 | -0.54 | - | -3.31 | - | - | - | - | - | -5.5(-0.67) | - | - | - | -2.28 | - | -0.7 | - | -1.36 |
| Tau mVC 2 | - | -0.55 | - | -4.92 | - | - | - | - | - | -5.1 | - | - | - | -2.03 | - | -0.89 | - | -0.99 |
| Tau mVC 3 | - | -0.46 | - | - | - | - | - | - | - | - | - | - | - | -2.04 | - | - | - | -0.56 |
| Tau mVC 4 | -0.42(-0.06) | -0.41 | - | - | - | - | 0(-0.16) | 0(-0.3) | - | - | - | - | -0.81(-0.26) | -1.69(-0.6) | - | - | -0.61 | -0.52 |
| Tau mVC 5 | - | -0.58 | - | -3.91 | - | - | - | - | - | -4.52 | - | - | - | -1.83 | - | -0.62 | - | -0.71 |

**Table S-9**. **Longitudinal association between the Montreal cognitive assessment (MoCA) battery and other cognitive scores applied to the OASIS participants and the multimodal variance component scores (mVC scores) extracted from the OASIS validation analysis**. Estimated difference in cognition between -2 and +2 standard deviations in mVC scores. Only significant mean differences (longitudinal annual cognitive decline for the same shift in mVC scores) after FDR correction at 0.05 significance level are presented (see methods). Poisson mixed effects regressions were used for estimation.

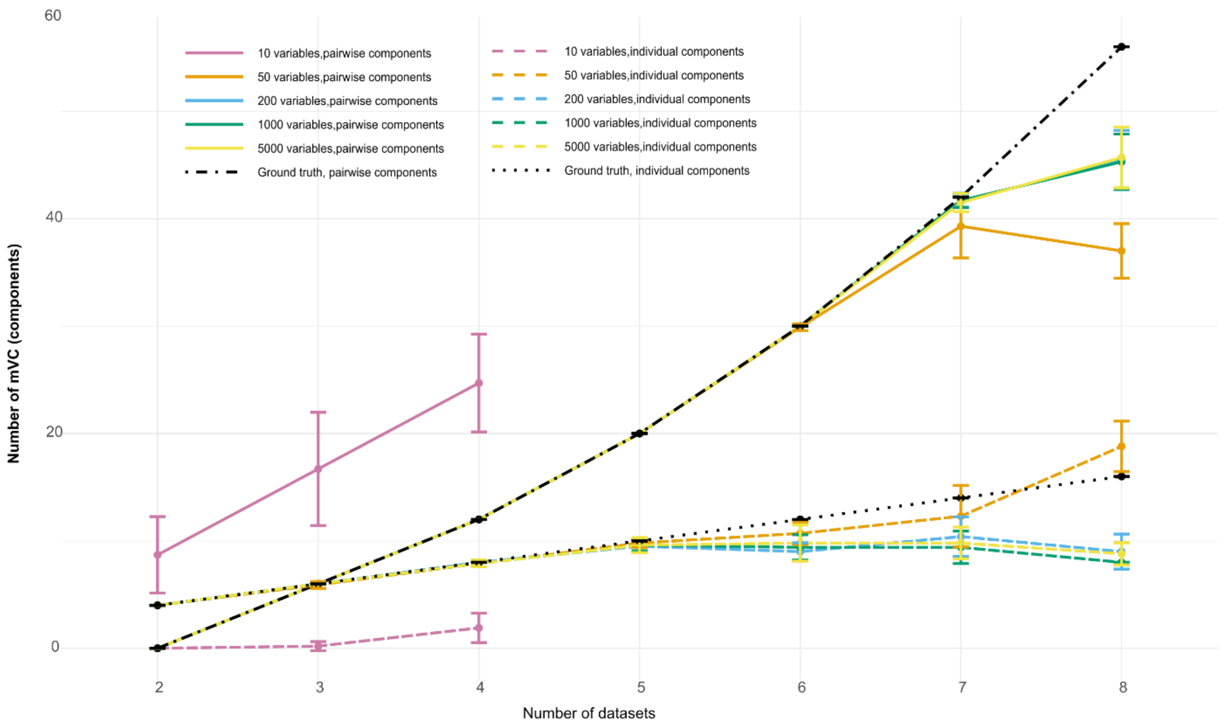


**Figure S-3. Estimated and simulated number of multimodal variance components (mVC) for multiview analysis with increasing number of datasets and variables**. The individual and pairwise mVC numbers are presented as mean and standard deviation extracted from 10 simulations with random seed. It can be seen the 50-5000 variable solutions had almost perfectly estimated number of mVCs until the 7-datasets solution. The 8-datasets solution had slightly reduced number of estimated mean mVCs suggesting that the model is able to accurately estimate pairwise mVCs for up to 7 datasets and with 50-5000 variables. The 10-variable within dataset solution had higher estimates than the ground truth suggesting that increasing number of datasets must be accompanied by increasing number of variables in each dataset. The joint mVCs are not presented here since they were estimated with no error apart from four combinations in the 10 and 50-variables solution. Specifically for the 10-variables and 2, 3 and 4-datasets solution, the ground truth was 4, 2, and 2 mVCs, while the mean estimated mVCs were 1.9, 1.2, and 0.9 respectively. Specifically for the 50-variables and 8-datasets solution, the ground truth was 2 mVCs and the mean estimated mVCs were 1.5.

| mVC type | Number of datasets | 50 variables | | | 200 variables | | | 1000 variables | | | Ground truth number of mVCs |
|---|---|---|---|---|---|---|---|---|---|---|---|
| | missing data | 1% | 10% | 30% | 1% | 10% | 30% | 1% | 10% | 30% | |
| Individual | 2 | 4(0) | 4(0) | 6.6(2.7) | 4(0) | 4(0) | 4(0) | 4(0) | 4(0) | 4(0) | 4 |
| | 6 | 10.8(1.5) | 12.4(1.8) | 21.6(1.7) | 9.2(1.2) | 9.1(1.3) | 7.5(1.2) | 10(1.2) | 9.2(1.3) | 6.4(0.7) | 12 |
| pairwise | 2 | 0(0) | 0(0) | 0(0) | 0(0) | 0(0) | 0(0) | 0(0) | 0(0) | 0(0) | 0 |
| | 6 | 29.9(0.) | 27.6(1.7) | 14.2(1.6) | 30(0) | 30(0) | 27.3(1.8) | 30(0) | 30(0) | 28(1.1) | 30 |
| joint | 2 | 4(0) | 4(0) | 3.8(0.4) | 4(0) | 4(0) | 4(0) | 4(0) | 4(0) | 4(0) | 4 |
| | 6 | 2(0) | 2(0) | 2(0) | 2(0) | 2(0) | 2(0) | 2(0) | 2(0) | 2(0) | 2 |

**Table S-10. Estimated number of multimodal variance components (mVC) when missing values are added to all the datasets.** The estimated number of individual, pairwise, and joint mVCs when missing values are introduced in the 2 and 6-dataset models are good for up to 10% missing values. For 30% missing data, individual and pairwise components are over or under-estimated for the 50, 200 and 1000-variable solutions. Results are presented as mean (standard deviation).

| mVC type | Number of datasets | 50 variables | | | 200 variables | | | 5000 variables | | | Ground truth number of mVCs |
|---|---|---|---|---|---|---|---|---|---|---|---|
| | missing data | 1% | 10% | 30% | 1% | 10% | 30% | 1% | 10% | 30% | |
| Individual | 2 | 4(0) | 4(0) | 4(0) | 4(0) | 4(0) | 4(0) | 4(0) | 4(0) | 4(0) | 4 |
| | 6 | 11.5(0.9) | 11.7(1) | 12(1.1) | 10.2(0.9) | 9.8(0.8) | 9.9(0.7) | 9.9(1.5) | 10.5(1.5) | 10.5(1) | 12 |
| pairwise | 2 | 0(0) | 0(0) | 0(0) | 0(0) | 0(0) | 0(0) | 0(0) | 0(0) | 0(0) | 0 |
| | 6 | 29.8(0.42) | 29.8(0.42) | 29.6(0.52) | 29.9(0.32) | 30(0) | 30(0) | 30(0) | 30(0) | 30(0) | 30 |
| joint | 2 | 4(0) | 4(0) | 4(0) | 4(0) | 4(0) | 4(0) | 4(0) | 4(0) | 4(0) | 4 |
| | 6 | 2(0) | 2(0) | 2(0) | 2(0) | 2(0) | 2(0) | 2(0) | 2(0) | 2(0) | 2 |

**Table S-11. Estimated number of multimodal variance components (mVC) when missing values are added to only one dataset.** The estimated number of individual, pairwise, and joint mVCs when missing values are introduced in only one dataset in the 2 and 6-dataset models are good for up to 30% missing values in some cases. For 30% missing data some individual mVCs are over or under for the 200 and 5000-variable solutions. Results are presented as mean (standard deviation).

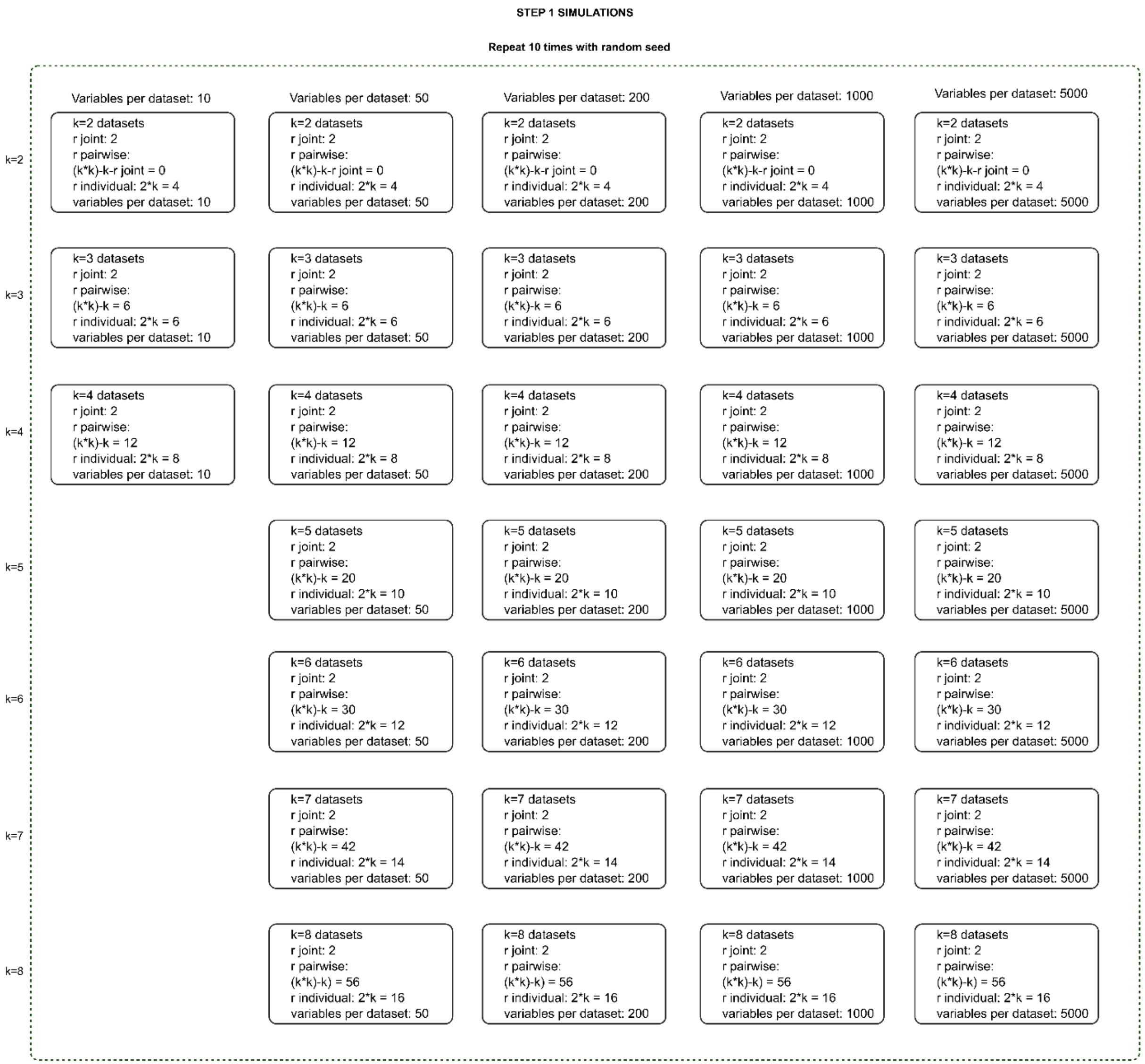


**Figure S-4.** Flowchart of simulations for the first batch of simulations. Specifications: samples= 300; signal to noise ratio= 1; loadings were simulated to be orthogonal.

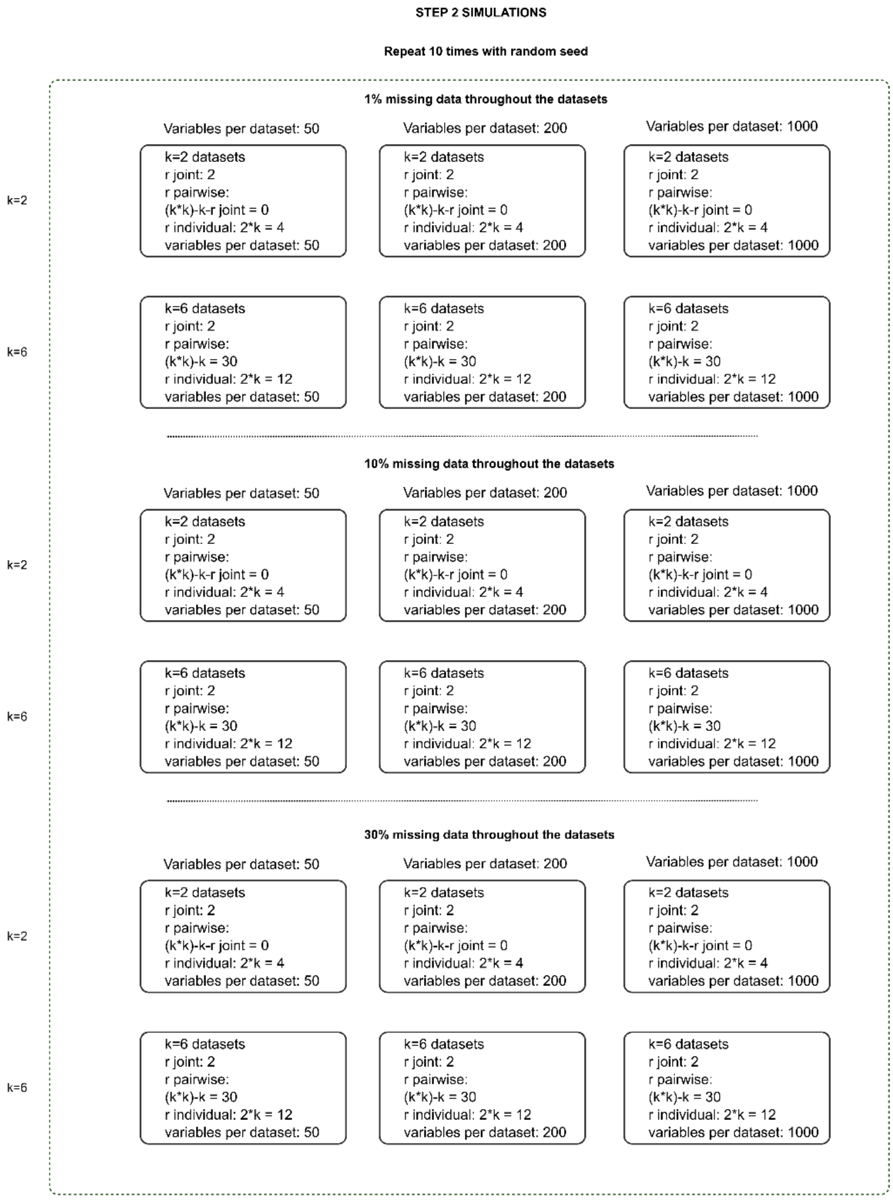


**Figure S-5.** Flowchart of simulations for the second and third batch of simulations. The third batch of simulations that included structured missingness in one of the datasets only, has the same specifications as the second batch but 5000 variable datasets are also tested. Specifications: samples= 300; signal to noise ratio= 1; loadings were simulated to be orthogonal.

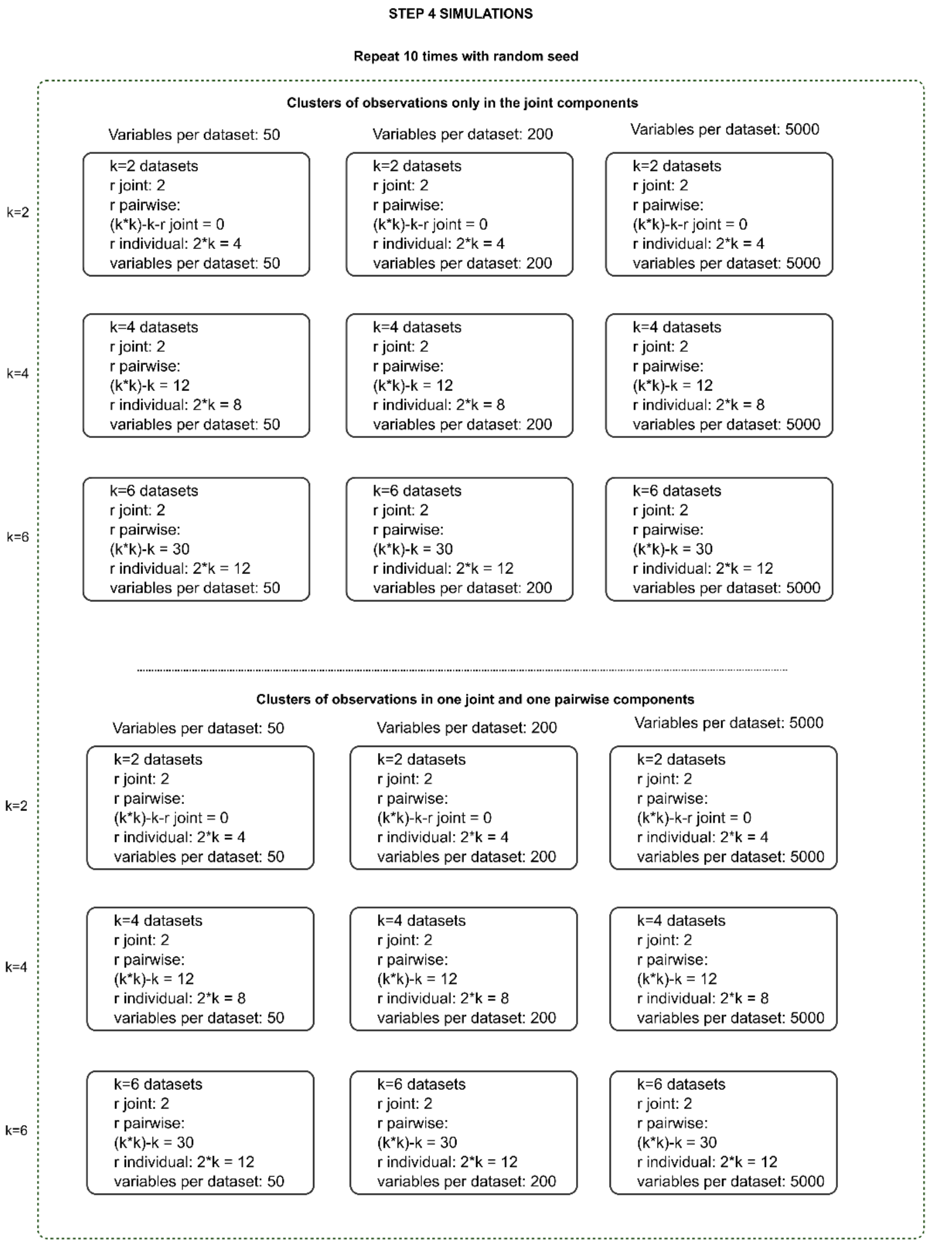


**Figure S-6.** Flowchart of simulations for the fourth batch of simulations. The fourth batch of simulations included clusters of samples in the datasets. Its has similar specifications with the third batch of simulations while clusters of samples are tested in one or two multimodal variance components. Specifications: samples= 320; signal to noise ratio= 1; loadings were simulated to be orthogonal.

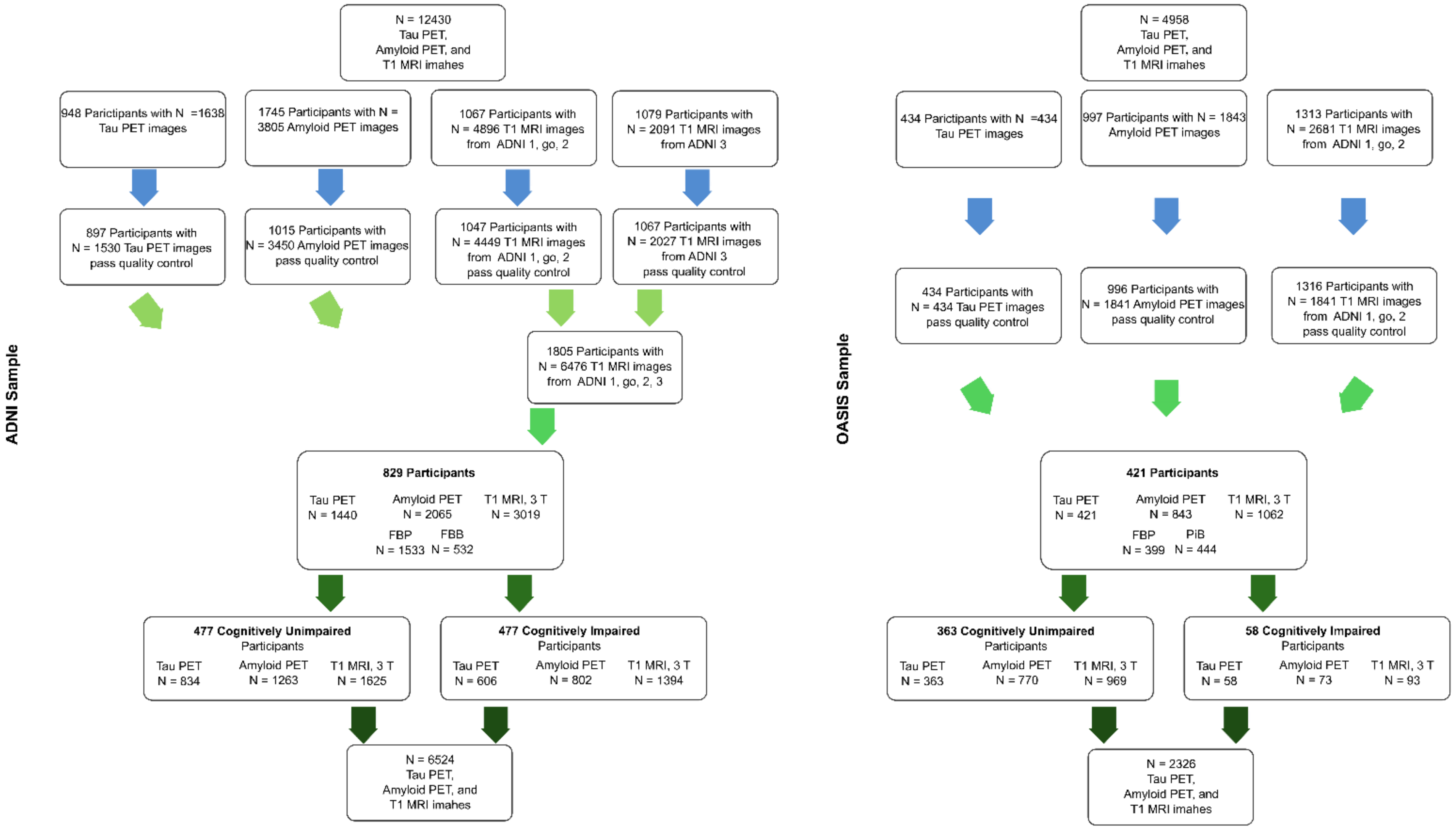


**Figure S-7. Subject and image selection in the ADNI and OASIS samples.**
